# Fatigue symptoms associated with COVID-19 in convalescent or recovered COVID-19 patients; a systematic review and meta-analysis

**DOI:** 10.1101/2021.04.23.21256006

**Authors:** Sanjay Rao, Tarek Benzouak, Sasha Gunpat, Rachel J. Burns, Tayyeb A. Tahir, Stephen Jolles, Steve Kisely

## Abstract

**Background:** The prevalence and prognosis of post-acute stage SARS-CoV-2 infection fatigue symptoms remain largely unknown.

**Aims:** We performed a systematic review to evaluate the prevalence of fatigue in post-recovery from SARS-CoV-2 infection.

**Method:** Medline, Embase, PsycINFO, CINAHL, Web of Science, Scopus, trial registries, Cochrane Central Register of Controlled Trials and Google Scholar were searched for studies on fatigue in samples that recovered from PCR diagnosed COVID-19. English, French and Spanish studies were included. Meta-analyses were conducted separately for each recruitment setting.

**Results:** We identified 41 studies with 9362 patients that recovered from COVID-19. Post-COVID-19 patients self-report of fatigue was higher compared to healthy controls (RR = 3.688, 95%CI [2.502, 5.436], p < 0.001). Over 50% of patients discharged from inpatient care reported symptoms of fatigue during the first (ER = 0.517, 95%CI [0.278, 0.749]) and second month following recovery (ER = 0.527, 95%CI [0.337, 0.709]). 10% of the community patients reported fatigue in the first-month post-recovery. Patient setting moderated the association between COVID-19 recovery and fatigue symptoms (R^2^ = 0.11, p < 0.001). Female patients recovering from COVID-19 had a greater self-report of fatigue (OR = 1.782, 95%CI [1.531, 2.870]). Patients recruited through social media had fatigue above 90% across multiple time points. Fatigue was highest in studies from Europe.

**Conclusion:** Fatigue is a symptom associated with functional challenges which could have economic and social impacts. Developing long-term planning for fatigue management amongst patients beyond the acute stages of SARS-CoV-2 infection is essential to optimizing patient care and public health outcomes. Further studies should examine the impact of sociodemographic, pandemic-related restrictions and pre-existing conditions on fatigue.

---

The first cases of SARS-CoV-2, the virus responsible for the COVID-19 disease, were reported in November 2019[1]. By March 2020, the SARS-CoV-2 outbreak was declared a pandemic by the World Health Organization (WHO)[2]. The acute presentation of SARS-COV-2 consists of both mental and physical health symptoms[3 4]. However even after resolution of the infection, some symptoms may persist for a significant period of time[5-7]. The terms ‘long Covid’, ‘long haul Covid’, and ‘Covid long hauler’ are frequently used in the media and scientific literature to capture the challenges of individuals suffering from such post-Covid syndromes[8-10]. The WHO has specifically recognized the occurrence of longer-term health effects of an acute SARS-COV-2 infection[8 11]. Thus far there are no clinical diagnostic criteria for such syndromes. Fatigue is the most frequent symptom described[12-15] followed by sleep difficulties, hair loss, olfactory dysfunction, cardiac palpitations, joint pain, negative changes to appetite, taste dysfunction, dizziness, diarrhoea or vomiting, chest pain, throat discomfort or difficulties swallowing, rash, myalgia, headaches and fever[12].

Chronic fatigue has been associated with previous infections with cytomegalovirus (CMV)[16], Epstein-Barr Virus (EBV)[17], Herpesvirus-6[18], Ross River virus[19], Dengue virus[20], MERS[21], and SARS-CoV-1[22]. However, the post viral fatigue syndromes cannot be explained by organ damage. The causes of post-viral symptoms during recovery stages are largely unknown. Longer-term changes in the immune system[20 23] or dysregulation of peripheral and autonomic nervous system functioning[24 25] have been implicated.

The US library of medicine defines fatigue as “fatigue is a feeling of weariness, tiredness, or lack of energy”[26]. Central to fatigue is the awareness of a decreased capacity for physical or mental function due to the presence of dysfunction in the availability, utilization and/or restoration of resources needed to perform a task[27]. Fatigue symptoms are usually identified through self-report and may be measured through fatigue rating instruments[28]. Many criteria have been devised to describe fatigue syndromes [29]. The Oxford Criteria (1991) for Chronic Fatigue Syndrome, includes a subtype of ‘post infectious fatigue syndrome (PIFS)’[30]. Furthermore, this criterion accepts the presence of mood symptoms although they are not necessary to diagnose Chronic Fatigue. Although a duration of 6 months or greater is included in some criteria guidelines, the 2011 International Consensus Criteria dropped the duration requirement[31]. Fatigue at rest and/or slow recovery from fatigue is highlighted across the different criteria [29].

Longer term COVID-19 syndromes can fall into more than one pattern[32 33], and the underlying causal mechanisms may be many. For instance, in certain individuals, cardiac function changes have been described in patients with symptoms of fatigue after contracting and recovering from acute SAR-Cov-2 infection[34 35]. Patients admitted to the ICU may also have more extensive lung damage[36] with fatigue as a symptom[37]. However, this explanation may not apply to the vast majority of cases of fatigue post COVID infection[38-40]. Similarly, although fatigue can be a symptom of depression, post viral fatigue cannot be fully explained by low mood[41].

In some countries, such as the UK, services to address the long-term effects of COVID-19 have been proposed[42 43] and are being established[44 45]. In this context, fatigue is an important symptom because it is associated with disability[46 47] and economic consequences[48 49]. Fatigue symptoms may also affect morale and predispose to other psychiatric conditions such as depression[50 51]. Various professional bodies have published initial guidelines for the management of such symptoms[52]. It is possible that even patients with mild forms of COVID-19 who do not receive inpatient care may still suffer long-term symptoms[53].

The current literature is unclear about the incidence and prevalence of such symptoms in individuals with a past diagnosis of SARS-Cov-2 and how long they persist. As the number of patients recovering from COVID-19 continues to rise, addressing long-term consequences, such as fatigue, are critical for optimizing the health outcomes of survivors.

This systematic review aims to address essential questions specific to post-Covid fatigue to inform and guide the evaluation and management of COVID-19 recovery. First, the study will provide a quantitative evaluation of post-Covid fatigue prevalence across different time points from cross-sectional and longitudinal studies. Where possible, the review will also examine associations with factors such as the severity of illness, type of patient population (e.g., inpatient vs outpatient), effect of gender, comorbidities and different definitions of recovery.

## Methods

This systematic review was conducted in accordance with the PRISMA [54] and MOOSE guidelines. The study protocol and MEDLINE search strategy were pre-registered in the Open Science Framework (i.e., osf.io/zu25b) on the 14th of September, 2020 and underwent full registration in the International Prospective Register of Systematic Reviews on the 18th of September, 2020 (i.e., [CRD42020209411]).

### Search strategy

We searched Medline, Embase, PsycINFO, CINAHL, Web of Science, Scopus, trial registries (i.e., NIH clinical trials registry, Cochrane Central Register of Controlled Trials and ISRCTN registry) and Google Scholar. Pre-print servers (MedRxiv and Psycharxiv) were also included in our search. The database search was undertaken on the 16th of September, 2020, the 1st of October, 2020, the 8th of November, 2020, and the 14th of February, 2021. The study selection process was applied to these new searches. A manual search of OpenGrey was further conducted to identify grey literature, and 62 authors with publications relating to COVID-19 patient follow-up were contacted with requests to share unpublished data relating to fatigue symptoms.

A combined set of MeSH and keywords associated with COVID-19 and fatigue were used to identify publications on fatigue in individuals recovered from acute COVID-19 infection, diagnosed with appropriate testing. No restrictions relating to study design, location, or language were imposed. The search strategy was synthesized by one of the authors (TB) and reviewed by a medical librarian. The strategy for each database is provided in Supplementary Material S-2. Only publications from 2019 onwards were considered. References from opinion publications relating to post-COVID fatigue were hand searched and screened. Study authors were contacted when PCR reports were ambiguous or for further data on the course of fatigue.

Rayyan (QCRI) software was used for screening and managing the abstracts. Title, abstract, and full-text screening were conducted by two independent reviewers (TB and SG), and a senior author (SR) resolved any screening discordance. Study selection criteria were applied to the retrieved full text articles independently by two reviewers (TB and SG). Any disagreements were resolved through a discussion between the reviewers and a senior author (SR).

### Selection Criteria

The eligibility criteria were: a) COVID-19 was diagnosed by Rapid Testing polymerase chain reaction test (RT-PCR) or Viral Antigen Test; b) Recovery was defined by a negative finding in one of these tests and/or clinical judgement; c) Fatigue was assessed on follow-up either through self-report, clinical interview or a fatigue specific scale.

The exclusion criteria were: a) COVID-19 status was screened through antibody testing (i.e., IgG and IgM) only; b) absence of a post-infection follow-up; c) the sample comprised solely of participants with specific medical conditions (e.g., Parkinson’s disease, Acquired Immunodeficiency Syndrome or transplant patients); d) the sample was comprised of participants below the age of 18; e) publication was in a language other than English, French or Spanish; or f) utilizing non-human methodology (i.e., lab simulation, in vitro or animal models).

As post-viral effects related to COVID-19 are an evolving challenge, several study designs were considered for evaluation. These included the following: cluster or non-cluster randomized controlled trials, controlled trials, and uncontrolled trials, cross-sectional, case-control and cohort studies.

### Data Extraction

Data extraction was initiated on the 20th of December, 2020, and completed on the 20th of February, 2021. Data extraction was conducted by reviewers (TB and SG) in consultation with a senior author (SR). Study authors were contacted if required data were missing from a publication. Descriptive data extracted in this systematic review included the author, year of publication, study location, COVID-19 diagnostics, participants’ mean age, study setting (i.e., clinical setting, general population), fatigue symptom report, study sample size, attrition, length of time between recovery and follow-up assessments of fatigue. For each report of post-viral fatigue, continuous data (i.e., mean and standard deviation) and categorical data (i.e., frequency data) relating to post-viral fatigue were extracted for meta-analysis.

### Assessment of Bias and Quality

Study quality assessments were completed by two reviewers (TB and SG). A third reviewer (SR) resolved any discrepancies. Risk of bias was assessed using several instruments, matching the appropriate scale to the study design. For non-randomized studies, the National Institute of Health Study Quality Assessment Tool for Observational Cohort and Cross-Sectional Studies was selected. For randomized studies, Cochrane’s ROB 2 was identified. The Joanna Briggs level of evidence scale for prognosis[55] was used for the overall strength of evidence.

### Data analysis

Comprehensive Meta-Analysis software (version 3) was used to conduct a meta-analysis of the data from the selected studies. Analyses were initiated on the 21st of February, 2021. A random-effects model was used for all statistical analyses. The primary outcome was the presence of fatigue. We calculated pooled prevalence (i.e., Event rate; ER) and two-tailed 95% confidence intervals in studies reporting post-viral fatigue on follow-up. Prevalence analyses were pooled by recruitment setting: inpatient, outpatient, mixed, registries, and social media or COVID-19 app-based settings. For each recruitment setting, meta-analyses were performed for studies that applied uniform measurements of fatigue (i.e., self-report and validated measurement tools) and in which follow-up was conducted within the same month post-recovery. We used Risk Ratio (RR) as an effect size metric in studies that compared patients who recovered from COVID-19 with a control group. Odds Ratios (OR) were also used where COVID-19 recovered patients were divided into groups based on the severity of the acute infection or type of hospital care received. ORs were calculated to compare patients with PCR test negative and those retested positive after recovery. ORs were also used for examining gender differences and estimating the effect of biomarkers. Meta-regressions were conducted on studies rated as fair or good quality. We first examined the moderating effect of self-reported fatigue compared to the use of fatigue rating scales. The majority of the studies assessed fatigue through self-reports. Data from these were used to examine the effect of time since recovery, PCR negative tests to confirm recovery, recruitment setting, average sample age, proportion of female participants, Diabetes Mellitus, COPD and hypertension. An analysis of event rate as a function of continent was further conducted, followed by a sub-group analysis of differences between continental regions.

Heterogeneity was tested using the Q statistic and reported in the form of a percentage of variation across studies (I^2^). A visual inspection of publication bias was conducted using a funnel plot and Egger’s linear regression modelling to statistically determine the presence of symmetry.

## Results

### Study Selection

The initial search of the literature identified 4384 abstracts. Four hundred ninety-eight abstracts were selected. Full-text articles of these abstracts were obtained, and study criteria were applied. Thirty-three published studies[6 12-15 32 33 38 40 41 56-78] and eight medRxiv preprints[53 79-85], met the inclusion criteria for this systematic review (Figure 1). The sample sizes ranged from 18[74] to 1655[12], with mean age across studies ranging from 32.3 (SD = 8.5)[33] to 67.1 (SD =11.6)[63]. The studies represented 18 countries with a total sample of 8825 individuals recovered from COVID-19 (Table 1). Diagnosis of COVID-19 was confirmed through Polymerase Chain Reaction (PCR) testing and PCR negative status used to confirm recovery in 16 studies (5190 cases). Twenty-five studies (4172 cases) defined recovery as hospital discharge or a specified number of days since the last positive PCR results. Twenty-six studies were from inpatient settings and five from outpatient settings, while four studies recruited through a mix of different clinical settings. Two studies recruited through patient registries, and one utilized a combination of two epidemiological datasets. Three studies reported recruitment through other methods such as social media (k = 1), a COVID-19 symptom app (k = 1) or a mix of online recruitment streams (k = 1). Thirty-one studies measured fatigue through symptom self-report and 10 studies utilized validated fatigue scales to evaluate fatigue rates within their samples.

**Table 1.**
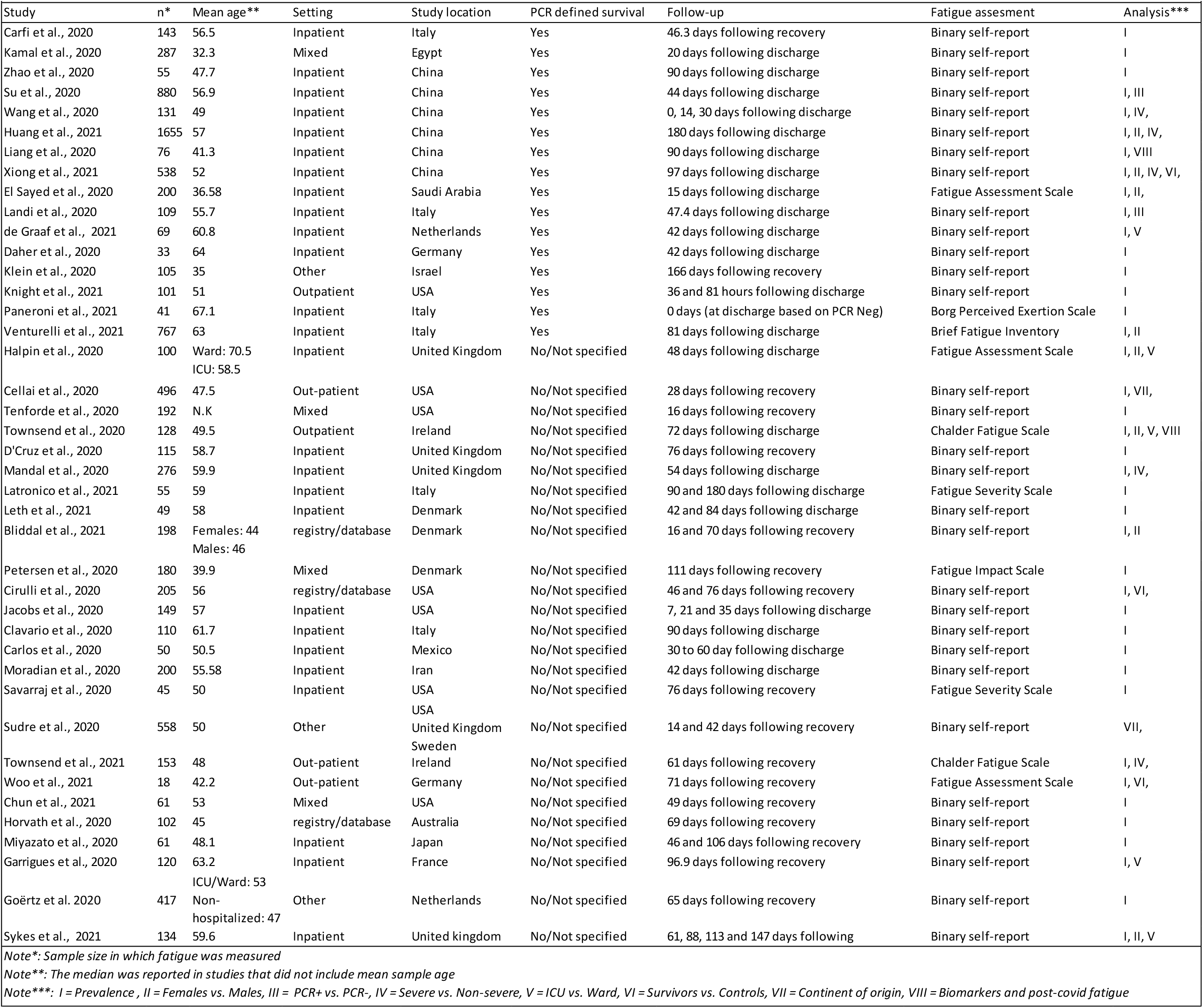
Characteristics of included studies

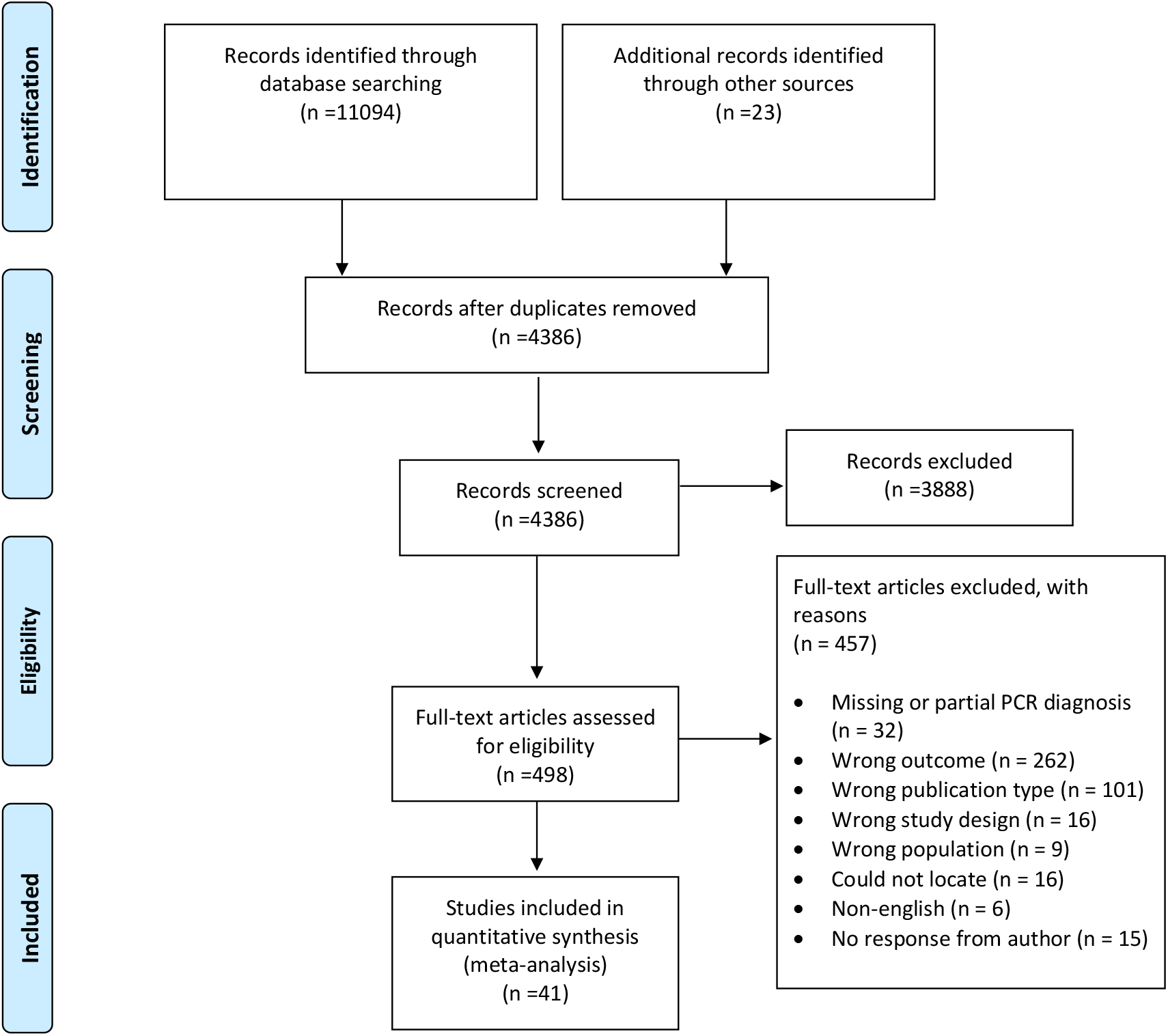

Five studies compared patients discharged from ICUs and hospital wards[14 32 40 60 77]. Five studies compared recovery from severe and non-severe experiences of SARS-CoV-2 infection[12 38 56 58 67]. Three studies compared COVID-19 recovered patients and healthy controls[58 74 81]. Eight studies compared residual fatigue in female and male survivors[12 14 40 41 53 58 64 77], and two compared patients who retested positive for SARS-Cov-2 on PCR test with those who retrained PCR negative status[59 79]. Finally, two studies evaluated the association between biomarkers and post-COVID fatigue[40 57].

Data were extracted from 41 studies and organized primarily according to the study setting and time intervals over which fatigue was assessed. Thirty-one (i.e., 31/41) studies used self-report of fatigue and ten used fatigue rating scales. The time intervals or temporal groups were: 0-30 days (1 month), 31-60 days (1-2 months), 61-90 days (2-3 months), 91-120 days (3-4 months), 121-150 days (4-5 months), and 151-180 days (5-6 months). Studies differed in the way recovery from COVID-19 was defined. Therefore, for each study, we chose one of the following approaches: PCR negative test, number of days after discharge from the hospital, and number of days since onset of the symptoms or PCR positive test. Fifteen studies defined their follow-up time as the number of days since symptom onset or PCR positive test results. In these studies, to account for the acute phase of COVID-19, day-0 of recovery was defined as fourteen days after follow-up initiation as this represents the recommended length of self-quarantine for patients with COVID-19[86 87]

Eight had repeated measure designs[53 56 62 68 69 71 76 81]. Where there were multiple assessments of fatigue within a temporal group, we calculated the mean of the fatigue scores or mean of the proportion of patients reporting fatigue.

### Quality assessment and Publication bias

Risk of bias assessment was conducted using the NIH Quality Assessment Tool for observational studies. Each study was assessed on the basis of its own reported details and guided by the NIH scores obtained. During bias quality assessment an emphasis was placed on sample recruitment characteristics, temporal aspects of follow-up, the use of statistical analyses, the presence of validated measures of fatigue outcomes and the use of PCR to define survival. Most studies were of fair quality (see S-6). The interrater reliability of the assessments was high (k = 0.8047, %agreement = 95.12%), and discrepancies were resolved through consensus. However, most studies were exploratory in nature rather than hypothesis driven. The absence of a clearly defined hypothesis, in addition to the absence of validated fatigue assessment tools, defining recovery without PCR testing and a lack of repeated follow-ups represented the most common limitations within included studies. Based on JBI methodology, the evidence for prognosis was level 3.a, and there was a wide range of study designs restricting definitive conclusions on quantifying fatigue post-recovery from COVID-19.

Publication bias was assessed through funnel plots and Egger’s regression. A low level of publication bias was observed when all studies were considered (see S-5; Egger’s bias = -3.049, 95%CI [-6.777, 0.680), t = 1.654, p = 0.106). No significant publication bias was found in studies with self-reported fatigue (Egger’s bias = -2.687, 95%CI [-7.445, 2.070), t = 1.154, p = 0.258). However, studies using fatigue scales showed significant publication bias (Egger’s bias = -3.797, 95%CI [-7.534, -0.059), t = 2.34, p = 0.047).

#### 1. Fatigue in COVID-19 recovered patients from inpatient settings

In the twenty-five studies with inpatient settings, the duration of follow-up was from 1-6 months. The majority of these studies assessed fatigue through self-reports (k = 20). 10 studies used PCR negative test as a proxy for recovery.

In Group 1 (<1 month), 3 out 5 studies used self-report of fatigue. There was a non-significant trend towards lower pooled prevalence of fatigue with the use of validated scales at 32% (k = 2, ER = 0.320, 95%CI [0.126, 0.607], I^2^ = 88.14%) compared to self-reports at 51.7% (k = 3, ER = 0.517, 95%CI [0.278, 0.749], I^2^ = 92.81%). However, the heterogeneity associated with these studies was found to be high. No meaningful comparison could be made between methods of defining recovery as 4 out of 5 studies used PCR negative testing to confirm recovery. With 12 studies, Group 2 (1-2 months) had the largest sample size and all studies used self-reports of fatigue. The pooled prevalence of fatigue was 52.7% (k = 7, ER = 0.527, 95%CI [0.337, 0.709], I^2^ = 96.07%; see figure 2a) in studies that did not use PCR negative test to confirm recovery compared to 49.3% (k = 7, ER = 0.493, 95%CI [0.204, 0.787], I^2^ = 98.36%; see figure 2b) in studies that used PCR negative test. This difference was nonsignificant (Q = 0.037, df = 1, p > 0.05) and there was considerable heterogeneity between studies.

**Figure 2a.**
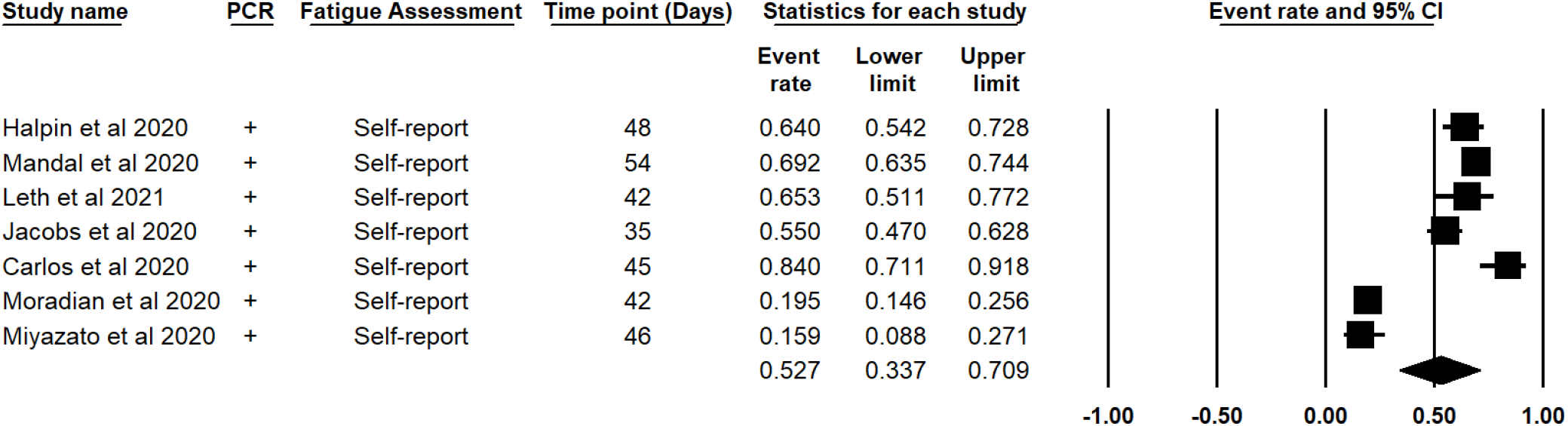
Prevalence of self-reported fatigue within the second month following inpatient recovery defined by clinical assessment rather than PCR negative testing.

**Figure 2b.**
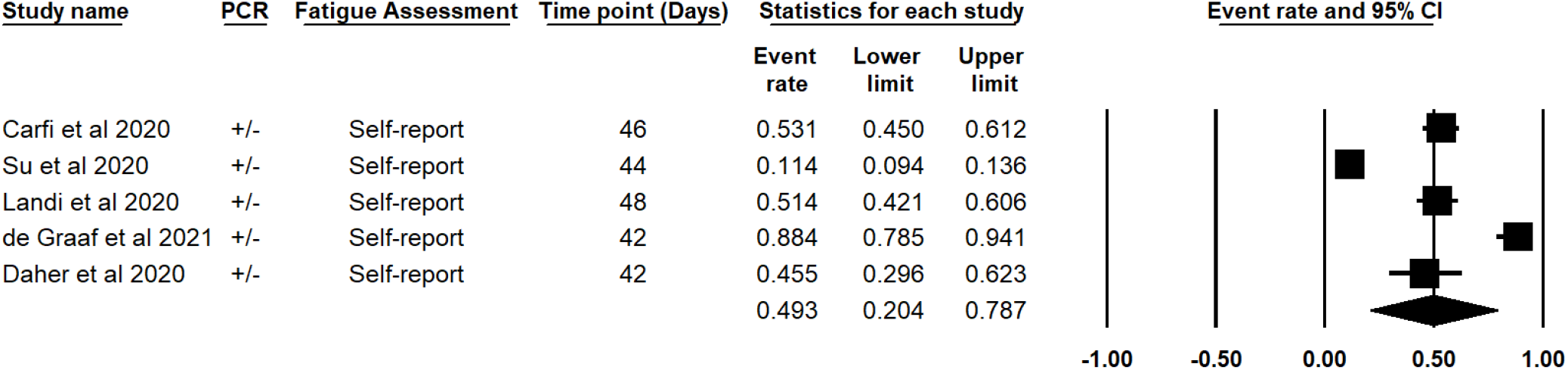
Prevalence of self-reported fatigue within the first month following inpatient recovery defined by PCR negative testing.

**Figure 2c.**
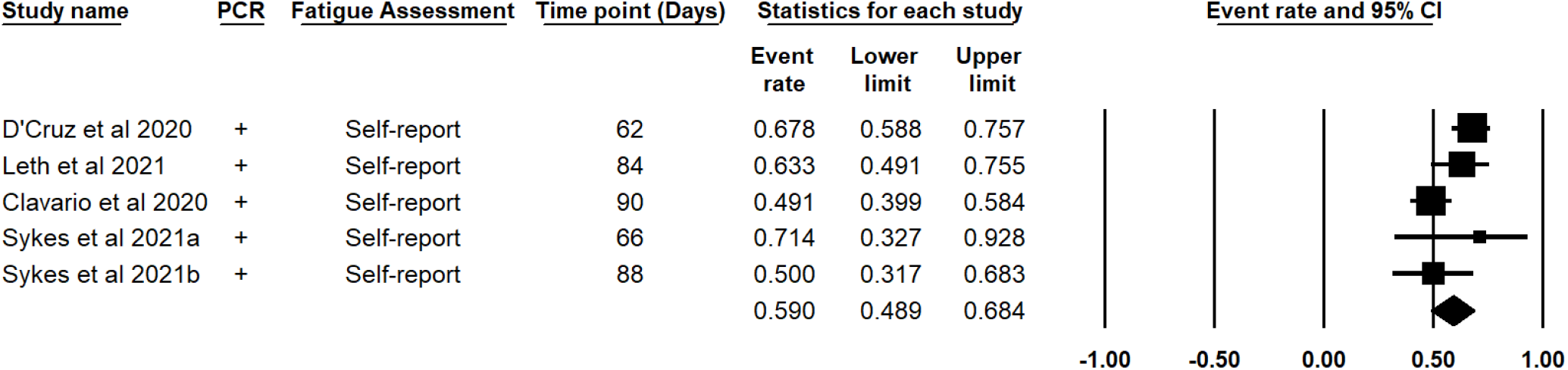
Prevalence of self-reported fatigue within the third month following inpatient recovery defined by clinical assessment rather than PCR negative testing.

**Figure 2d.**
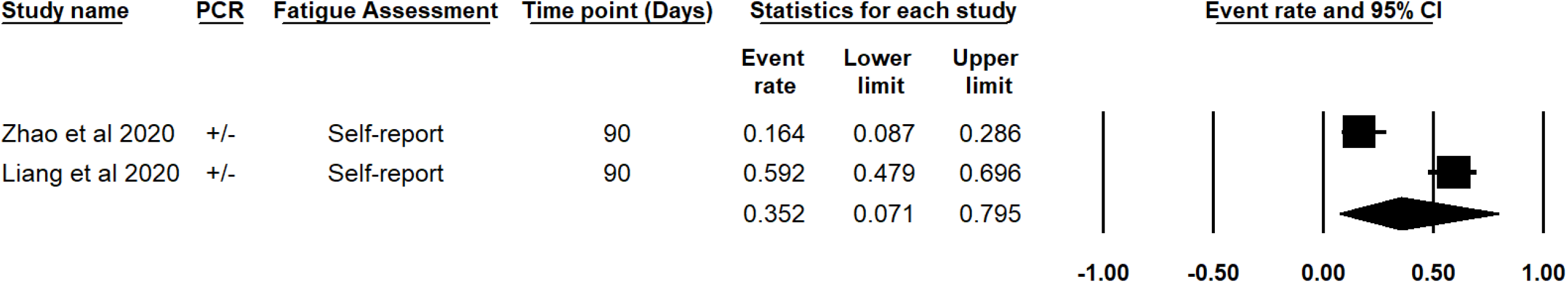
Prevalence of self-reported fatigue within the third month following inpatient recovery defined by PCR negative testing.

**Figure 2e.**
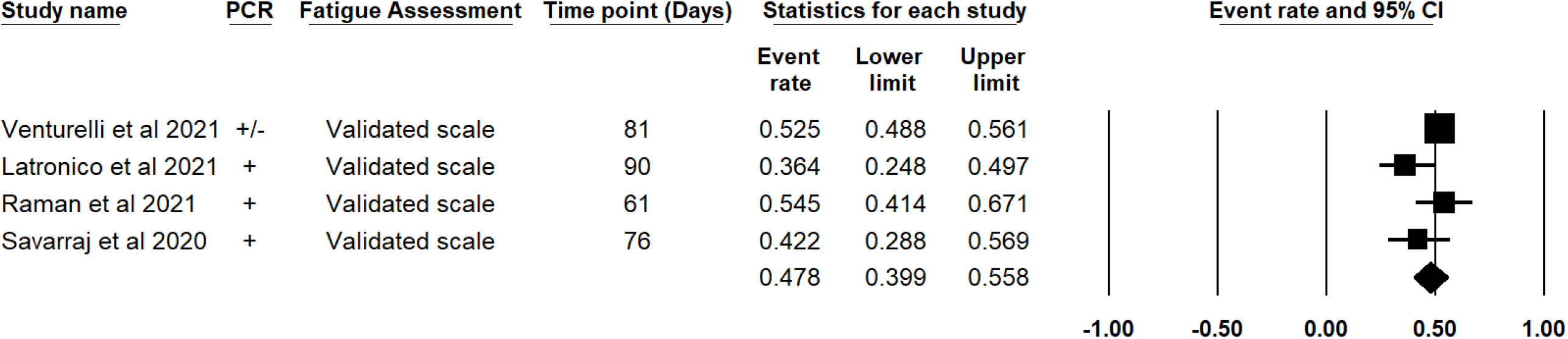
Prevalence of fatigue within the third following inpatient recovery, in which fatigue was measured using validated fatigue scales.

In Group 3 (2-3 months), 11 studies assessed fatigue. Seven of these studies used self-reports of fatigue and 4 used fatigue scales. Self-reported fatigue (k = 7, ER = 0.525, 95%CI [0.392, 0.653], I^2^ = 83.85%) was higher in studies that did not use PCR negative test to define recovery (k=5, ER = 0.590, 95%CI [0.189, 0.684], I^2^ = 58.83%; see figure 2c) compared to those that did not (k=2, ER = 0.352, 95%CI [0.071, 0.795], I^2^ = 95.34%; see figure 2d) but this did not reach significance (p > 0.05). Pooled prevalence of fatigue measured with scales was 47.8% (k = 4, ER = 0.478, 95%CI [0.399, 0.558], I^2^ = 56.29%; see figure 2e) with moderate heterogeneity between studies. No meaningful comparison could be made between methods of defining recovery within these studies as only 1 study used PCR negative test to confirm recovery. The pooled prevalence of fatigue measured through scales was lower than the analysis of self-reported fatigue, this difference was nonsignificant (Q = 0.424, df = 1, p > 0.05).

In the 3-4 month period (Group 4), there were four studies that elicited self-report of fatigue. The prevalence of self-reported fatigue was 30.1% (k = 4, ER = 0.301, 95%CI [0.171, 0.427], I^2^ = 92.98%). Only 1 study used the PCR negative test result to identify recovery and had a fatigue prevalence of 28.3% (n = 538, 95%CI [0.246, 322]). The three smaller studies that did not use PCR negative test status for recovery, had a wide difference in fatigue prevalence with a 29.8% pooled prevalence of fatigue (k = 3, ER = 0.298, 95%CI [0.117, 0.577], I^2^ = 93.48%).

Group 5 (4-5 months) had only one study[77] which assessed self-reported fatigue. The prevalence of fatigue within their sample was found to be 39.1% (k = 1, ER = 0.391, 95%CI [0.218, 0.598]). Notably, this study did not use PCR to define recovery.

Two studies followed up recovered patients beyond 6 months (Group 6). Huang and colleagues measured fatigue through self-report and defined recovery through PCR negative test. Within their sample, 1038 (62.7%) reported fatigue out of a total of 1655 participants (95%CI [0.604, 0.650]). In contrast, Latronico et al. also evaluated fatigue 180 days following discharge and assessed fatigue using the Fatigue Severity Scale rather than dichotomous self-reports. This single point estimate found a 35.6% (95%CI [0.231, 0.504]) prevalence of residual fatigue in COVID-19 recovered cases; this assessment did not use the PCR test to confirm recovery from COVID-19. However, the study conducted by Latronico et al. had a lower precision of estimate, likely due to a small sample size of 55 participants.

Overall, inpatient reports included data for the first 6 months following recovery from COVID-19. The total prevalence of self-reported fatigue was 47.1% (k = 24, ER = 0.471, 95%CI [0.367, 0.578], I^2^ = 97.27%). Similarly, fatigue as measured by validated assessment resulted in a prevalence of 43.3% (k = 6, ER = 0.433, 95%CI [0.352, 0.516], I^2^ = 77.04%). However, each analysis was found to have a high level of heterogeneity and were found to not significantly differ from each other (Q = 0.278, df = 1, p = 0.598).

#### 2. *Fatigue in COVID-19 recovered patients from out*patient settings

Five studies measured fatigue outcomes in COVID-19 in the first, second, and third month following recovery.

In Group 1 (<1 month) the larger study by Cellai et al. did not use PCR negative test for recovery and reported fatigue in only 3.4% (n = 496, 95%CI [0.021, 0.054]) of patients. However, this may have been a result of the study design. Only a small number of the total sample were assessed for symptoms after 3 weeks. The Knight et al. study established recovery through PCR test negative findings. Of the sample, 34.7% (n = 101, 95%CI [0.260, 0.444]) and 28.4% (n = 95, 95%CI [0.203, 0.383]) reported fatigue during follow-ups conducted 36 days and 81 days following recovery status.

The only study of fatigue between 1-2 months (Group 2) was Townsend et al. This study evaluated the presence of post-COVID fatigue. The prevalence of fatigue within their sample was 52.3% (n = 128, 95%CI [0.437, 0.608]). Fatigue was measured using the Chalder Fatigue Scale, but recovery was not defined using PCR testing.

Two studies assessed fatigue using a validated scale within 2-3 months (Group 3). Woo et al. had a smaller sample size of 18 and a very wide time interval over which patients were assessed for symptoms. The prevalence of fatigue was 16.7% over 20-105 days. In contrast, Townsend et al. (2021) reported fatigue in 47.7% of their cohort (n = 153, 95%CI [0.399, 0.556]). Neither study used PCR negative testing to confirm recovery, and the data from these studies were not pooled due to significant differences in the duration of follow-up.

Overall, outpatient reports of self-reported fatigue included data for the first month and validated assessments ranged from 58 to 85 days following recovery from COVID-19. The total prevalence of self-reported fatigue was determined to be 11.3% (k = 2, ER = 0.113, 95%CI [0.010, 0.611], I^2^ = 98.34%). Fatigue as measured by validated assessment resulted in a prevalence of 44.9% (k = 3, ER = 0.449, 95%CI [0.329, 0.575], I^2^ = 70.57%) within the first 3 months. However, both analyses had a high level of heterogeneity. Although a 33.6% difference was observed between overall self-reports of fatigue and fatigue as measured by validated scales, the low number of studies resulted in a lack of precision in determining significant differences (Q = 2.565, df = 1, p = 0.11).

#### 3. *Fatigue in COVID-19* recovered patients from mixed settings

Four studies evaluated fatigue in patients that recovered from COVID-19 in mixed settings. Two studies examined self-reported fatigue during the first month (Group 1) following recovery and consisted of patients that recovered from in-and outpatient care, and non-hospitalized cases. Data from these studies were not pooled due to significant differences in their follow-up design. First, 72.8% (n = 287, 95%CI [0.674, 0.777]) of the cohort in a study conducted by Kamal et al. reported experiencing fatigue. This study used PCR to define recovery from COVID-19. Another study conducted within this timeframe by Tenforde et al. found a prevalence of 71% when considering only respondents to telephone follow-up; however, this prevalence dropped to 35.4% when considering the full sample in which non-respondents are included.

One study was found to report fatigue during the second month of recovery within a mixed sample (i.e., hospital and non-hospital care). Within their sample, the prevalence of fatigue was 29.5% (n = 61, 95%CI [0.194, 0.421]).

Peterson et al. used the Fatigue Impact Scale over an average follow-up of 111 days, although this covered a wide range of 45-215 days. The fatigue reported within their cohort was lower than in other studies as the prevalence was 8.3% (n = 180, 95%CI [0.051, 0.134]); this difference is most likely affected by the study methodology, which included an extensive interquartile follow-up range.

The overall analysis could only be computed for studies in which fatigue was self-reported and ranged from 2 to 63 days following recovery. This resulted in a fatigue prevalence of 46.3% (k = 3, ER = 0.463, 95%CI [0.202, 0.746], I^2^ = 97.42%). Datapoints were found to be highly heterogeneous.

#### 4. Fatigue in COVID-19 recovered subjects recruited from general registry and secondary data

Three studies analyzed self-report data from non-hospitalized patient registries (k = 2) and non-selective databases (k = 1). None defined recovery by PCR, and all measured fatigue using symptom self-report. Pooling two data points within the first month (Group 1) of recovery resulted in a prevalence of 9.7% (k = 2, ER = 0.097, 95%CI [0.031, 0.262], I^2^ = 91.10%; see S-4).

One study by Cirulli and colleagues assessed fatigue during the second month of recovery. Within their sample, 6.6% (n = 152, 95%CI [0.036, 0.118]) reported the presence of fatigue symptoms 46 days following recovery.

All three studies evaluated fatigue symptoms during the third month of recovery, in which pooling point estimates resulted in an 8.2% (k = 3, ER = 0.082, 95%CI [0.034, 0.188], I^2^ = 80.75%; see S-4) prevalence of fatigue. Figures from all different time points in this study were substantially lower than post-COVID fatigue in the hospital population.

#### 5. Fatigue in COVID-19 recovered subjects recruited using social media or tech-based apps

Three studies recruited individuals that recovered from COVID-19 using social media (k = 1), a COVID-19 related app (k = 1), and mixed online recruitment methods (k = 1).

A repeated measures study conducted by Sudre et al. used a mobile symptom tracking COVID-19 app. Within their sample, fatigue was reported by 97.7% of 558 recovered cases (ER = 0.977, 95%CI [0.960, 0.986]) during follow-up participation 28 days post-positive PCR test results. Furthermore, when assessing fatigue in 189 individuals that recovered from COVID-19 56 days following PCR diagnosis of COVID-19, the prevalence was found to be 96.8% (k = 1, ER = 0.968, 95%CI [0.931, 0.986]). The study aimed to identify symptoms associated with long-COVID; fatigue was found to be the most prevalent symptom within individuals reporting persistent symptoms following COVID-19 recovery. Fatigue during the first week of diagnosis was the strongest predictor of fatigue reports during a follow-up 28 days post-diagnosis fatigue (k = 1, OR=2.83 95%CI [2.09;3.83]). This study remains ongoing and is now assessing over 4182 incident cases of COVID-19.

Pooling results from Sudre et al., and a follow-up conducted by Goërtz et al., demonstrated a presence of fatigue of 93.6% (k = 2, ER = 0.936, 95%CI [0.655, 0.991], I^2^ = 97.77%) during follow ups between 56 days to 79 days following initial symptom onset. However, both studies did not use PCR to define recovery and heterogeneity within the analysis was found to be high.

Klein et al. recruited their sample using social media 5 to 6 months (Group 5) post recovery and employed a snowball recruitment method. The prevalence of self-reported fatigue was found to be 21.90% (K = 1, ER = 0.219, 95%CI [0.150, 0.308]) 180 days post-PCR diagnosis.

Pooled analysis of all studies in which recruitment was completed through social media, app base or COVID-19 website determined a 79.7% prevalence of fatigue within the first 166 days following recovery (k = 3, ER = 0.797, 95%CI [0.264, 0.977], I^2^ = 99.00%). Heterogeneity was found to be high.

#### 6. Overall reported fatigue amongst patients recovered from COVID-19

Considering all studies, irrespective of setting or temporal characteristics, self-reported fatigue was found to have a prevalence rate of 42% (k = 32, ER = 0.420, 95%CI [0.319, 0.529], I^2^ = 98.09%) within the first six months of recovery. In contrast, fatigue prevalence as defined by validated assessment was found to be present in 34.9% of recovered cases (k = 9, ER = 0.349, 95%CI [0.255, 0.457], I^2^ = 92.07%). The difference between self-reported fatigue and fatigue as defined by validated assessment did not reach the precision necessary to determine a significant difference (Q = 0.656, df = 1, p = 0.418).

Only two studies examined fatigue severity using a continuous scale. The first by El Sayed et al., reported a mean score of 40.81 ± 5.75 on the Fatigue Assessment Scale during a follow-up 15 days post-negative PCR results. The second conducted by Townsend et al. reported a mean score of 15.8 ± 5.75 on the Chalder Fatigue Scale during a follow-up conducted 72 days post-discharge. Although their use of different scales did not permit pooled prevalence of severity analyses, pooling of these studies resulted in a non-significant association between days since recovery and fatigue symptom intensity (k = 2, r = 0.430, 95%CI [-0.419, 0.878], z = 0.994, p = 0.320; I^2^ = 99.405%).

#### 7. Comparing fatigue in COVID-19 recovered patients with healthy controls

Three studies compared individuals recovered from COVID-19 with healthy controls. The studies collected participants from inpatients[58], outpatients[74] and an epidemiological database[81]. Pooled analysis of fatigue data was conducted for the period between 76-97 days, and fatigue assessment for all studies was in the form of self-report. COVID-19 patients were found to have a 3.688 increase in relative risk of fatigue outcomes compared to non-COVID-19 exposed groups (k = 3, RR = 3.688, 95%CI [2.502, 5.436], z = 6.592, p < 0.001; I2 = 0%; see S-4). The heterogeneity within these studies was low.

One study[81] assessed fatigue over 3-time points in which the COVID-19 recovered group had an increased relative risk of fatigue across time points when compared to healthy controls. These follow-ups were conducted 16 days (n = 4021, RR = 4.451, 95%CI [2.341, 8.464], p < 0.001), 46 days (n = 3221, RR = 4.696, 95%CI [2.406, 9.163], p < 0.001) and 76 days (n = 2821, RR = 5.530, 95%CI [2.746, 11.136], p < 0.001) following recovery. However, this is likely explained by higher attrition in the post-COVID-19 group compared to the control group. Notably, there were not enough data points to conduct a meta-analysis.

#### 8. Fatigue in Covid-19 PCR negative vs PCR positive patients

Two studies had data from discharged inpatients comparing those who retested PCR positive to those who were persistently PCR negative. No significant difference was found between patients identified as recovered from COVID-19 and patients retested positive for COVID-19 (k = 2, RR = 0.806, 95%CI [0.476, 1.363], p = 0.420, I^2^ = 53.02%; see S-4).

#### 9. Gender and fatigue reports amongst COVID-19 recovered patients

Eight studies had data comparing gender within COVID-19 recovered patients from both inpatient[12 14 41 58 64 77] and outpatient[40] settings and epidemiological datasets[53]. Female patients were more likely to self-report fatigue between 84-180 days of follow-up (Group 3-6) in pooled data from 4 studies (k = 4, OR =1.782, 95%CI [1.531, 2.870], z = 3.366, p = 0.001, I^2^ = 52.51%; see S-4). Sensitivity analysis in which only studies from Groups 2 to 3 were included further replicated the observed gender effects (k = 3, OR = 2.096, 95%CI [1.531, 2.870], z = 4.620, p < 0.001, I^2^ = 0%), and heterogeneity for this analysis was low. However, this difference disappeared when fatigue was assessed using rating scales (see S-4) in two studies across two time points of 12-48 days (k = 2, OR = 1.254, 95%CI [0.0.273, 5.756], p = 0.771, I^2^ = 89.92%), and 72 to 81 days post-COVID-19 recovery (k = 2, OR = 1.503, 95%CI [0.456, 4.951], p = 0.503, I^2^ = 90.63%).

#### 10. Fatigue in Severe vs. Non-Severe SARS-CoV-2 patients

Two studies similar in design and duration of follow-ups, compared recovered individuals with severe COVID-19 and non-severe COVID-19 related illnesses. When examining outcomes 54 to 97 days following recovery, individuals recovering from severe cases of COVID-19 did not significantly differ from one another with respect to fatigue (OR = 1.344, 95%CI [0.958, 1.886], z = 1.711, p = 0.087, I^2^ = 0%; see S-4) and other chronic symptoms[58 67]. Data from 2 other studies, which could not be pooled due to differences in study design, had similar findings; in which no difference was observed during the first month following PCR negative findings (OR = 1.181, 95%CI [0.262, 5.326], p = 0.829), and 75 days following discharge in which fatigue was assessed using the Chalder Fatigue Scale (OR = 0.711, 95%CI [0.397, 1.274], p = 0.252). Similarly, no significant difference was detected when evaluating evidence comparing patients discharged from the ICU and hospital wards during a follow-up within the second and third month of recovery (k = 2, OR = 0.991, 95%CI [0.332, 2.960], z = -0.017, p = 0.987, I^2^ = 60.23%; see S-4) in which fatigue was assessed using validated scales, and second to fourth month in which fatigue was self-reported (k = 3, OR = 1.024, 95%CI [0.566, 1.853], z = 0.078, p = 0.938, I^2^= 0%; see S-4).

In contrast, Huang et al. found within a sample of 1733 COVID-19 recovered cases that had previously received care in the form of a high-flow nasal cannula for oxygen therapy, non-invasive ventilation or invasive mechanical ventilation were 2.73 times (OR = 2.725, 95%CI [1.694, 4.381]) more likely to experience post-COVID fatigue when compared to patients that had previously received no supplementary oxygen. Perhaps the divergent findings may be linked to the different ways in which severity is defined. A more objective measure of severity was utilized in the study, which showed a difference between the severe and non-severe groups.

#### 11. Biomarkers and Post-COVID fatigue

Two studies examined immunological markers and their association with fatigue in patients that recovered from COVID-19. These studies were not pooled due to design differences in fatigue assessments. With regard to the two-point estimates, Liang et al. found no significant effect on fatigue outcomes when examining the role of CD3 (p > 0.05), CD4 (p > 0.05), CD8 (p > 0.05) lymphocytes. Furthermore, both pro-inflammatory IL-6 (p > 0.05), and CRP (p > 0.05) were nonsignificant predictors of fatigue 90 days following PCR negative testing. This finding was mirrored by Townsend et al. in which lymphocytes (p > 0.05), IL-6 (p > 0.05) and CRP (p > 0.05) were nonsignificant predictors of fatigue 72 days post-hospital discharge. However, patients recovered from COVID-19 within the Liang et al.’s sample were found to be 94.76 times more likely to experience fatigue for each unit increase in serum troponin-I (95%CI [24.935, 360.149], p < 0.001), suggesting cardiovascular implications. However, the current review did not have the necessary data points to evaluate this meta-analytically.

#### 12. Country of origin and fatigue

Extracted studies provided sufficient data points for comparison of self-reported fatigue in Asia, North-America, and Europe within the first six months of recovery from COVID-19. Self-reported fatigue was dependent on the continent of origin when comparing data points from Europe, North-America, and Asia (Q = 10.62, df = 2, p = 0.005). Europe had the highest levels of self-reported fatigue post-recovery from COVID-19 (K = 13, ER = 0.593, 95%CI [0.431, 0.736], I^2^= 94.60%), followed by North-America (K = 7, ER =0.300, 95%CI [0.149, 0.510], I^2^ = 97.58%) and Asia (K = 9, ER = 0.225,95%CI [0.116, 0.392], I^2^ = 98.83%). Heterogeneity was high within the continental data points. However, multivariate meta-regression determined that the significant variance explained by continental differences was robust as it remained significant (k = 29, Q = 15.24, df = 2, p < 0.001) when holding recruitment setting constant. Within this meta-regression, only the comparisons between Europe and Asia were significant in the context of controlling for sample setting. Asian studies were associated with a 1.84% decrease in self-reported prevalence of fatigue when compared to European studies (β = -1.840, 95%CI[-2.808, -0.870], z = -3.72, p < 0.001).

#### 13. Moderator analysis

Meta-regression was employed to examine the explained variance between point estimates and only included studies assessed to be of fair or good quality. Studies with repeated measures were excluded from the moderation analysis. When considering the first six months following COVID-19 recovery, the comparison between self-reported fatigue and fatigue assessed using validated scales did not significantly differ regarding COVID-19 prevalence (k = 28, β = -0.147, 95%CI[-1.044, 0.751], Q = 0.10, df = 1, p = 0.749).

All further moderation analyses were carried out using self-reported fatigue. The association between COVID-19 recovery and fatigue was not found to be dependent on the use of PCR negative testing to define recovery (k = 21, β = 0.406, 95%CI[-0.622, 1.435], Q = 0.60, df = 1, p = 0.439), average sample age (k = 19, β = 0.035, 95%CI[-0.035, 0.106], z = 0.98, p = 0.325), sample gender proportions (k = 21, β = -3.683, 95%CI[-7.576, 0.211], z = -1.85, p = 0.064), proportions of Diabetes Mellitus (k = 20, β = 3.283, 95%CI[-0.826, 7.391], z = 1.57, p = 0.117) or the sample prevalence of COPD (k = 12, β = 10.421, 95%CI[-13.550, 34.393], z = 0.85, p = 0.394). However, the association between COVID-19 recovery and fatigue was found to be dependent on recruitment setting (k = 21, R^2^ = 0.11, Q = 16.72, df = 3, p < 0.001); in which, both outpatient (β = -3.377, 95%CI[-5.550, -1.204], z = -3.05, p = 0.002) and samples from patient registry (β = -3.535, 95%CI[-5.945, -1.125], z = -2.87, p = 0.004) were found to be associated with decreased prevalence in self-reported fatigue. Furthermore, hypertension was found to be associated with COVID-19 recovery and fatigue (k = 19, R^2^ = 0.02, β = 2.777, 95%CI[0.148, 5.406], z = 2.07, p = 0.038). Overall, when considering 6 months following COVID-19 recovery, the length since recovery was not a significant mechanism of change for the association between COVID-19 exposure and post-recovery fatigue outcomes (K = 21, β = 0.003, 95%CI[-0.012, 0.017], z = 0.37, p = 0.708), even when holding patient setting constant (k = 21, β = -0.002, 95%CI[-0.016, 0.013], z = -0.21, p = 0.837).

## Discussion

This systematic review shows that self-reported fatigue after recovery from COVID-19 infection can last up to 6 months based on the longest duration studies. Patients in the post-acute stage of COVID-19 were 3.7 times more at risk for onset of fatigue compared to healthy controls. The prevalence of fatigue within the first six months of recovery was 42% for self-reported fatigue and 34.9% for cases where fatigue was measured using a validated scale. Between 30% to 60% of inpatient and outpatient treated patients reported fatigue. However, at the population level, the proportion of COVID-19 recovered patients suffering fatigue may be lower (i.e., < 10%). The highest proportion of fatigue (i.e., up to 90%) was amongst those persons who were recruited through social media and COVID-19 apps. This could be the effect of self-selection as those with persistent symptoms are more likely to use these channels to report on their health. The use of negative PCR tests to confirm recovery did not influence fatigue. Even when the test is negative, non-viral shedding may continue to occur[88], which may explain this finding.

There was an insufficient number of studies comparing self-report to the assessment of fatigue through rating scales. However, in female patients, fatigue was higher compared to males but only on self-report and not on assessments using rating scales. The severity of COVID-19 did not appear to moderate the expression of fatigue. This may be a result of how disease severity is defined (e.g., based on admission to general wards or to ICU), which could be dependent on multiple factors, including local admission protocols. A more precise definition of severity used in at least one study did show an increase in self-reported fatigue in those deemed more unwell.

Surprisingly, age did not have an effect on self-report or rating scale scores of fatigue. Since mortality is higher in older adults, the survivors may have fewer symptoms overall. There is an effect of continent of origin (see S-7), but it is possible that this may be due to reliance on self-report rather than the use of more objective fatigue measurement instruments. Reinfection measured through PCR positive seroconversion did not appear to influence the proportion of people reporting fatigue. There are initial findings of links with biomarkers; however, this will require additional studies to confirm persistent associations.

### Limitations and Strengths

To the best of our knowledge, this is the first systematic review examining fatigue post-recovery from COVID-19 across a wide range of individuals and settings spanning several countries. However, the findings were affected by factors such as significant heterogeneity in study design, duration, population, method of assessment of fatigue. In addition, our search was restricted to articles in English, French, and Spanish. Although it remains very common for international studies to be published in English, some Asian specific databases (e.g., China National Knowledge Infrastructure database) were found to be inaccessible to our researcher team. Inclusion of such language specific databases could have added further insight to the current analysis.

Notably, social isolation and population demographics, such as patient ethnicity, can affect physical[89] and mental health[90], and have been determined to be associated with different prognoses in the context of COVID-19[91-93]. Data relating to fatigue risk as a function of ethnicity within socially diverse populations (e.g., United States) was not reported within the available studies examining fatigue in the context of COVID-19 recovery. The inclusion of such data would have added insight on potential associations between fatigue and racial disparities amongst COVID-19 survivors. The sudden restrictions imposed on the majority of the world’s population may also impact the expression of physical symptoms such as fatigue. Our review could not separate the influence of such factors due to data limitations in which the absence of appropriate controls was present.

We were also unable to examine fatigue because of inconsistency in how the various studies documented or defined chronic diseases. This would require access to individual patient data. Causality remains unknown as the evidence does not yet provide the group comparisons required to differentiate between mechanisms such as COVID-19 or other factors associated with the pandemic. For instance, pandemic-related restrictions or decreased physical activity could have played a role in reported fatigue incidence. However, more granular data of lifestyle changes and psychological responses to the pandemic is required to address these implications within the context of COVID-19 recovery.

The study highlights that clinical populations are at risk for persistent fatigue. This could inform how advice is given to those who have been treated for SARS-Cov-2 infection in the hospital or outpatient settings. We did not have information to analyse the effect of different waves of the infection.

### Future research and application to clinical practice

Health policy and leadership must prepare for a longer-term management plan for COVID-19 as there are significant needs beyond recovery from the acute infection. Fatigue, along with other symptoms, may also affect the ability to function and work. This may have a significant economic impact as well as an impact on the lives of significant others in the patients’ lives [49]. Future research should focus on more objective assessments of fatigue and standardized follow-up of post-COVID-19 patients. In the interest of public health, it should be possible to share anonymized individual data of persistent post-COVID-19 symptoms. New variants of coronavirus may have a different profile of fatigue and this warrants further research. Finally, more research is needed into the pathogenesis of what may be a range of long-term syndromes.

## Data Availability

Please contact Dr. Sanjay Rao for availability of data

## Author contributions

S.R., T.T., S.K., S.J., and T.B. contributed to the design and conception of the study. S.R., S.G., and T.B. were involved in screening the texts for inclusion in the review. S.G. and T.B. extracted the data. T.B. and S.R. led the analysis and interpretation of data. S.R. and T.B. led the manuscript’s write-up, and T.T. contributed to the manuscript write-up. S.R. and T.B. contributed to the analysis and interpretation of the synthesis. All other authors provided a critical review of the manuscript and assisted with revising the manuscript. T.B., coordinated and S.R., supervised the systematic review. All authors approved the final manuscript for submission.

