## Supplementary material for "Fatigue symptoms associated with COVID-19 in convalescent or recovered COVID-19 patients; a systematic review and meta-analysis": S-2

### OVID was used for MEDLINE, EMBASE, PSYCHINFO and Cochrane databases

1. coronavirus infections/ or coronaviridae/ or Coronaviridae Infections/

2. coronavirus/ or exp betacoronavirus/

3. (19nCoV or 2019-nCoV or 2019nCoV or corona virus 2 or coronavirus* 2 or corona* 2019 or COV 2 or COVID19* or COVID-19 or COVID or covid-19 or HCoV-19 or ncov* or nCov-2019 or novel corona* or SARS-COV-2 or SARSCOV-2 or SARSCOV2 or severe acute respiratory syndrome?).ti,ab.

4. ((new or novel or "19" or "2019" or Wuhan or Hubei or China or Chinese) adj3 (betacoronavirus* or beta coronavirus* or CoV or HCoV)).ti,ab.

5. (coronavirus* or corona virus* or covid* or "long covid*" or (long adj1 haul adj1 Covid) or (covid adj1 long adj1 haulers)).ti,ab.

6. ((Wuhan or Hubei) adj5 pneumonia).ti,ab.

7. or/1-6

8. exp Fatigue/ or Fatigue.ti,ab.

9. Mental Fatigue/

10. Fatigue Syndrome, Chronic/

11. Fibromyalgia/ or fibromyalgia.ti,ab.

12. Exhaust*.ti,ab.

13. Weary.ti,ab.

14. Weariness.ti,ab.

15. Letharg*.ti,ab.

16. (myalgic adj1 encephalomyelitis).ti,ab.

17. lassitude.ti,ab.

18. tiredness.ti,ab.

19. 8 or 9 or 10 or 11 or 12 or 13 or 14 or 15 or 16 or 17 or 18

20. 7 and 19

21. limit 20 to yr="2019 -Current"

### CINAHL

1. (MH "Coronavirus Infections")

OR (MH "Coronaviridae")

OR (MH "Coronaviridae Infections")

OR (MH "Coronavirus")

OR (TI "betacoronavirus" OR AB "betacoronavirus")

OR (TI "(19nCoV or 2019-nCoV or 2019nCoV or corona virus 2 or coronavirus* 2 or corona* 2019 or COV 2 or COVID19* or COVID-19 or COVID or covid-19 or HCoV-19 or ncov* or nCov-2019 or novel corona* or SARS-COV-2 or SARSCOV-2 or SARSCOV2 or severe acute respiratory syndrome?)" OR AB "(19nCoV or 2019-nCoV or 2019nCoV or corona virus 2 or coronavirus* 2 or corona* 2019 or COV 2 or COVID19* or COVID-19 or COVID or covid-19 or HCoV-19 or ncov* or nCov-2019 or novel corona* or SARS-COV-2 or SARSCOV-2 or SARSCOV2 or severe acute respiratory syndrome?)")

OR (TI "(coronavirus* or corona virus* or covid*)" OR AB "(coronavirus* or corona virus* or covid*)")

OR (TI “("long N1 covid*" or (long N1 haul N1 Covid) or (covid N1 long N1 haulers)”

OR (TI "((Wuhan or Hubei) N5 pneumonia)" OR AB "((Wuhan or Hubei) N5 pneumonia)")

AND

2. (MH "Fatigue+") OR (MH "Fatigue Syndrome, Chronic") OR (MH "Mental Fatigue")

OR (TI "Fatigue" OR AB "Fatigue")

OR (MH "Fibromyalgia")

OR (TI "Fibromyalgia" OR AB "Fibromyalgia")

OR (TI "Exhaust*" OR AB "Exhaust*")

OR (TI "Weary" OR AB " Weary")

OR (TI "Weariness" OR AB " Weariness")

OR (TI "Letharg*" OR AB "Letharg*")

OR (TI "myalgic N1 encephalomyelitis" OR AB "myalgic N1 encephalomyelitis")

OR (TI " lassitude" OR AB "lassitude")

OR (TI "tiredness" OR AB "tiredness")

### Scopus

TITLE-ABS-KEY ( coronavirus AND infections ) OR TITLE-ABS-KEY ( coronaviridae ) OR TITLE-ABS-KEY ( coronaviridae AND infections ) OR TITLE-ABS-KEY ( coronavirus ) OR TITLE-ABS-KEY ( betacoronavirus ) OR TITLE-ABS-KEY ( 19ncov ) OR TITLE-ABS-KEY ( 2019-ncov ) OR TITLE-ABS-KEY ( 2019ncov ) OR TITLE-ABS-KEY ( "corona virus 2" ) OR TITLE-ABS-KEY ( "coronavirus* 2" ) OR TITLE-ABS-KEY ( "corona* 2019" ) OR TITLE-ABS-KEY ( "COV 2" ) OR TITLE-ABS-KEY ( covid19* ) OR TITLE-ABS-KEY ( covid-19 ) OR TITLE-ABS-KEY ( covid ) OR TITLE-ABS-KEY ( covid-19 ) OR TITLE-ABS-KEY ( hcov-19 ) OR TITLE-ABS-KEY ( ncov* ) OR TITLE-ABS-KEY ( ncov-2019 ) OR TITLE-ABS-KEY ( "novel corona*" ) OR TITLE-ABS-KEY ( sars-cov-2 ) OR TITLE-ABS-KEY ( sarscov-2 ) OR TITLE-ABS-KEY ( sarscov2 ) OR TITLE-ABS-KEY ( "severe acute respiratory syndrome?" ) OR TITLE-ABS-KEY ( ( ( new OR novel OR "19" OR "2019" OR wuhan OR hubei OR china OR chinese ) PRE/3 ( betacoronavirus* OR "beta coronavirus*" OR cov OR hcov ) ) ) OR TITLE-ABS-KEY ( coronavirus* ) OR TITLE-ABS-KEY ( "corona virus*" ) OR TITLE-ABS-KEY ( covid* ) OR TITLE-ABS-KEY ( ( ( wuhan OR hubei ) PRE/5 pneumonia ) ) OR TITLE-ABS-KEY (long PRE/1 Covid) OR TITLE-ABS-KEY (long PRE/1 haul PRE/1 covid) OR  TITLE-ABS-KEY (covid PRE/1 long PRE/1 hauler) AND  (TITLE-ABS-KEY(Fatigue) OR TITLE-ABS-KEY(Mental Fatigue) OR TITLE-ABS-KEY(Fatigue Syndrome,Chronic) OR TITLE-ABS-KEY(Fibromyalgia) OR TITLE-ABS-KEY(Exhaust*) OR TITLE-ABS-KEY(Weary) OR TITLE-ABS-KEY(Weariness) OR TITLE-ABS-KEY(Letharg*) OR TITLE-ABS-KEY("myalgic PRE/1 encephalomyelitis") OR TITLE-ABS-KEY(lassitude) OR TITLE-ABS-KEY(tiredness))

### WEB OF SCIENCE

(TS=("Coronaviridae infection" OR "Coronaviridae" OR "Coronaviridae infection") OR TI=(coronavirus) OR AB=(coronavirus)  OR TS=(Betacoronavirus') OR TI=(19nCoV or 2019-nCoV or 2019nCoV or “corona virus 2” or (coronavirus* NEAR/1 2) or corona* NEAR/1 2019 or COV 2 or COVID19* or COVID-19 or COVID or covid-19 or HCoV-19 or ncov* or nCov-2019 or novel corona* or SARS-COV-2 or SARSCOV-2 or SARSCOV2 or “severe acute respiratory” NEAR/1 syndrome?) OR AB=(19nCoV or 2019-nCoV or 2019nCoV or “corona virus 2” or (coronavirus* NEAR/1 2) or corona* NEAR/1 2019 or COV 2 or COVID19* or COVID-19 or COVID or covid-19 or HCoV-19 or ncov* or nCov-2019 or novel corona* or SARS-COV-2 or SARSCOV-2 or SARSCOV2 or “severe acute respiratory” NEAR/1 syndrome?) OR TI=((new or novel or "19" or "2019" or Wuhan or Hubei or China or Chinese) NEAR/3 (betacoronavirus* or beta NEAR/1 coronavirus* or CoV or HCoV)) OR AB=((new or novel or "19" or "2019" or Wuhan or Hubei or China or Chinese) NEAR/3 (betacoronavirus* or beta NEAR/1 coronavirus* or CoV or HCoV)) OR TI=(coronavirus* or corona NEAR/1 virus* or covid*) OR AB=(coronavirus* or corona NEAR/1 virus* or covid*) OR TI=((Wuhan or Hubei) NEAR/5 pneumonia) OR AB=((Wuhan or Hubei) NEAR/5 pneumonia)) OR TI=((coronavirus* or corona virus* or covid* or long NEAR/1 covid* or (long NEAR/1 haul NEAR/1 Covid) or (covid NEAR/1 long NEAR/1 haulers))) OR AB=(coronavirus* or corona virus* or covid* or "long covid*" or (long adj1 haul adj1 Covid) or (covid adj1 long adj1 haulers))

AND

TS=(fatigue OR "Chronic fatigue syndrome" OR Fibromyalgia OR Mental Fatigue) OR TI=(fatigue OR "chronic fatigue syndrome" OR fibromyalgia OR exhaust* OR Weary OR Weariness OR Letharg* OR (myalgic NEAR/1 encephalomyelitis) OR lassitude OR tiredness) OR AB=(fatigue OR "chronic fatigue syndrome" OR fibromyalgia OR exhaust* OR Weary OR Weariness OR Letharg* OR (myalgic NEAR/1 encephalomyelitis) OR lassitude OR tiredness)

### Google scholar

("Coronaviridae infection" OR "Coronaviridae" OR "Coronaviridae infection" OR coronavirus OR coronavirus OR Betacoronavirus OR 19nCoV or 2019-nCoV or 2019nCoV or “corona virus 2” or “coronavirus 2” or “corona 2019” or COV 2 or COVID19 or COVID-19 or COVID or covid-19 or HCoV-19 or ncov or nCov-2019 or novel corona or SARS-COV-2 or SARSCOV-2 or SARSCOV2 or “severe acute respiratory syndrome” OR Wuhan or Hubei or China or Chinese betacoronavirus or coronavirus or CoV or HCoV OR coronavirus or “corona virus” or covid OR Wuhan or Hubei AND pneumonia) AND (fatigue OR "Chronic fatigue syndrome" OR Fibromyalgia OR Mental Fatigue OR fatigue OR "chronic fatigue syndrome" OR fibromyalgia OR exhaust OR Weary OR Weariness OR Letharg* OR myalgic encephalomyelitis OR lassitude OR tiredness)

***Search returned 1 paper, restructured search to enhance sensitivity.***

The final search for google scholar was:

COVID-19 AND FATIGUE AND ("POST-VIRAL") AND restrict date to from 2019

### MedRxiv (Major word count limit)

***Major wordcount limit, restructured search***

(SARS-Cov-2 OR COVID-19) AND ("chronic Fatigue" or Fibromyalgia or "persistent fatigue") AND (Persistent or Post-viral)

### ISRCTN registry

("Coronaviridae infection" OR "Coronaviridae" OR "Coronaviridae infection" OR coronavirus OR coronavirus OR Betacoronavirus OR 19nCoV or 2019-nCoV or 2019nCoV or “corona virus 2” or “coronavirus 2” or “corona 2019” or COV 2 or COVID19 or COVID-19 or COVID or covid-19 or HCoV-19 or ncov or nCov-2019 or novel corona or SARS-COV-2 or SARSCOV-2 or SARSCOV2 or “severe acute respiratory syndrome” OR Wuhan or Hubei or China or Chinese betacoronavirus or coronavirus or CoV or HCoV OR coronavirus or “corona virus” or covid OR Wuhan or Hubei AND pneumonia) AND (fatigue OR "Chronic fatigue syndrome" OR Fibromyalgia OR Mental Fatigue OR fatigue OR "chronic fatigue syndrome" OR fibromyalgia OR exhaust OR Weary OR Weariness OR Letharg* OR myalgic encephalomyelitis OR lassitude OR tiredness)

***Search returned 0 paper, restructured search to enhance sensitivity.***

The final search for the ISRCTN registry was:

(Covid-19 OR SARS-COV-2 ) AND fatigue

### PsyAxiv

("Coronaviridae infection" OR "Coronaviridae" OR "Coronaviridae infection" OR coronavirus OR coronavirus OR Betacoronavirus OR 19nCoV or 2019-nCoV or 2019nCoV or “corona virus 2” or “coronavirus 2” or “corona 2019” or COV 2 or COVID19 or COVID-19 or COVID or covid-19 or HCoV-19 or ncov or nCov-2019 or novel corona or SARS-COV-2 or SARSCOV-2 or SARSCOV2 or “severe acute respiratory syndrome” OR Wuhan or Hubei or China or Chinese betacoronavirus or coronavirus or CoV or HCoV OR coronavirus or “corona virus” or covid OR Wuhan or Hubei AND pneumonia)

AND

(fatigue OR "Chronic fatigue syndrome" OR Fibromyalgia OR Mental Fatigue OR fatigue OR "chronic fatigue syndrome" OR fibromyalgia OR exhaust OR Weary OR Weariness OR Letharg* OR myalgic encephalomyelitis OR lassitude OR tiredness)

***Search returned 1 paper, restructured search to enhance sensitivity.***

The final search for the PsyAxiv was:

(SARS-Cov-2 OR COVID-19) AND ("chronic Fatigue" or Fibromyalgia or "fatigue")

### https://clinicaltrials.gov/

("Coronaviridae infection" OR "Coronaviridae" OR "Coronaviridae infection" OR coronavirus OR coronavirus OR Betacoronavirus OR 19nCoV or 2019-nCoV or 2019nCoV or “corona virus 2” or “coronavirus 2” or “corona 2019” or COV 2 or COVID19 or COVID-19 or COVID or covid-19 or HCoV-19 or ncov or nCov-2019 or novel corona or SARS-COV-2 or SARSCOV-2 or SARSCOV2 or “severe acute respiratory syndrome” OR Wuhan or Hubei or China or Chinese betacoronavirus or coronavirus or CoV or HCoV OR coronavirus or “corona virus” or covid OR Wuhan or Hubei AND pneumonia) AND (fatigue OR "Chronic fatigue syndrome" OR Fibromyalgia OR Mental Fatigue OR fatigue OR "chronic fatigue syndrome" OR fibromyalgia OR exhaust OR Weary OR Weariness OR Letharg* OR myalgic encephalomyelitis OR lassitude OR tiredness)

***Search returned 0 paper, restructured search to enhance sensitivity.***

(SARS-Cov-2 OR COVID-19) AND ("chronic Fatigue" or Fibromyalgia or "fatigue")

***Search returned 0 paper, restructured search to enhance sensitivity.***

The final search for the ISRCTN registry was:

"fatigue" AND (Post-viral OR longterm OR recovery)
