## Supplementary material for "Fatigue symptoms associated with COVID-19 in convalescent or recovered COVID-19 patients; a systematic review and meta-analysis": S-3

Supplementary material 3 (S-3). List of included studies

1. Carfì, A., Bernabei, R., & Landi, F. (2020). Persistent symptoms in patients after acute COVID-19. Jama, 324(6), 603-605.
2. Kamal, M., Abo Omirah, M., Hussein, A., & Saeed, H. (2020). Assessment and characterisation of post-COVID-19 manifestations. International Journal of Clinical Practice, e13746.
3. Zhao, Y. M., Shang, Y. M., Song, W. B., Li, Q. Q., Xie, H., Xu, Q. F., ... & Luo, H. (2020). Follow-up study of the pulmonary function and related physiological characteristics of COVID-19 survivors three months after recovery. EClinicalMedicine, 25, 100463.
4. Su, Y., Zhu, L. S., Gao, Y., Li, Y., Xiong, Z., Hu, B., ... & Cheng, F. (2020). Clinical characteristics of Covid-19 patients with re-positive test results: an observational study. medRxiv.
5. Wang, X., Xu, H., Jiang, H., Wang, L., Lu, C., Wei, X., ... & Xu, S. (2020). The Clinical Features and Outcomes of Discharged Coronavirus Disease 2019 Patients: A Prospective Cohort Study. QJM: An International Journal of Medicine.
6. Huang, C., Huang, L., Wang, Y., Li, X., Ren, L., Gu, X., ... & Cao, B. (2021). 6-month consequences of COVID-19 in patients discharged from hospital: a cohort study. The Lancet.
7. Liang, L., Yang, B., Jiang, N., Fu, W., He, X., Zhou, Y., ... & Wang, X. (2020). Three-Month Follow-Up Study of Survivors of Coronavirus Disease 2019 after Discharge. Journal of Korean medical science, 35(47).
8. Xiong, Q., Xu, M., Li, J., Liu, Y., Zhang, J., Xu, Y., & Dong, W. (2020). Clinical sequelae of COVID-19 survivors in Wuhan, China: a single-centre longitudinal study. Clinical Microbiology and Infection, 27(1), 89-95.
9. El Sayed, S., Shokry, D., & Gomaa, S. M. (2020). Post-COVID-19 fatigue and anhedonia: A cross-sectional study and their correlation to post-recovery period. Neuropsychopharmacology Reports.
10. Landi, F., Carfì, A., Benvenuto, F., Brandi, V., Ciciarello, F., Monaco, M. R. L., ... & Team, P. A. C. (2020). Predictive Factors for a New Positive Nasopharyngeal Swab Among Patients Recovered From COVID-19. American Journal of Preventive Medicine, 60(1), 13-19.
11. de Graaf, M. A., Antoni, M. L., ter Kuile, M. M., Arbous, M. S., Duinisveld, A. J. F., Feltkamp, M. C. W., ... & Roukens, A. H. E. (2021). Short-term outpatient follow-up of COVID-19 patients: A multidisciplinary approach. EClinicalMedicine, 100731.
12. Daher, A., Balfanz, P., Cornelissen, C., Müller, A., Bergs, I., Marx, N., ... & Müller, T. (2020). Follow up of patients with severe coronavirus disease 2019 (COVID-19): Pulmonary and extrapulmonary disease sequelae. Respiratory medicine, 174, 106197.
13. Klein, H., Asseo, K., Karni, N., Benjamini, Y., Nir-Paz, R., Muszkat, M., ... & Niv, M. Y. (2020). Onset, duration, and persistence of taste and smell changes and other COVID-19 symptoms: longitudinal study in Israeli patients. medRxiv.
14. Knight, D., Downes, K., Munipalli, B., Halkar, M. G., Logvinov, I. I., Speicher, L. L., & Hines, S. L. Symptoms and Clinical Outcomes of Coronavirus Disease 2019 in the Outpatient Setting. SN Comprehensive Clinical Medicine, 1-8.
15. Paneroni, M., Simonelli, C., Saleri, M., Bertacchini, L., Venturelli, M., Troosters, T., ... & Vitacca, M. (2021). Muscle strength and physical performance in patients without previous disabilities recovering from COVID-19 pneumonia. American Journal of Physical Medicine & Rehabilitation, 100(2), 105-109.

16. Venturelli, S., Benatti, S. V., Casati, M., Binda, F., Zuglian, G., Imeri, G., ... & Rizzi, M. (2021). Surviving COVID-19 in Bergamo Province: a post-acute outpatient re-evaluation. Epidemiology & Infection, 1-25.
17. Halpin, S. J., McIvor, C., Whyatt, G., Adams, A., Harvey, O., McLean, L., ... & Collins, T. (2020). Postdischarge symptoms and rehabilitation needs in survivors of COVID-19 infection: A cross-sectional evaluation. Journal of medical virology.
18. Cellai, M., & O'Keefe, J. B. (2020, October). Characterization of prolonged COVID-19 symptoms in an outpatient telemedicine clinic. In Open forum infectious diseases (Vol. 7, No. 10, p. ofaa420). US: Oxford University Press.
19. Tenforde, M. W., Kim, S. S., Lindsell, C. J., Rose, E. B., Shapiro, N. I., Files, D. C., ... & Gong, M. N. (2020). Symptom duration and risk factors for delayed return to usual health among outpatients with COVID-19 in a multistate health care systems network—United States, March–June 2020. Morbidity and Mortality Weekly Report, 69(30), 993.
20. Townsend, L., Dyer, A. H., Jones, K., Dunne, J., Mooney, A., Gaffney, F., ... & Sugrue, J. A. (2020). Persistent fatigue following SARS-CoV-2 infection is common and independent of severity of initial infection. PloS one, 15(11), e0240784.
21. D'Cruz, R. F., Waller, M. D., Perrin, F., Periselneris, J., Norton, S., Smith, L. J., ... & Madula, R. (2020). Chest radiography is a poor predictor of respiratory symptoms and functional impairment in survivors of severe COVID-19 pneumonia. ERJ Open Research.
22. Mandal, S., Barnett, J., Brill, S. E., Brown, J. S., Denneny, E. K., Hare, S. S., ... & Hurst, J. R. (2020). 'Long-COVID': a cross-sectional study of persisting symptoms, biomarker and imaging abnormalities following hospitalisation for COVID-19. Thorax.
23. Latronico, N., Peli, E., Rodella, F., Novelli, M. P., Rasulo, F. A., Piva, S., & Center, L. O. T. O. (2020). Six-Month Outcome in Survivors of COVID-19 Associated Acute Respiratory Distress Syndrome. Six-Month Outcome in Survivors of COVID-19 Associated Acute Respiratory Distress Syndrome.
24. Leth, S., Gunst, J. D., Mathiasen, V. D., Hansen, K. S., Søgaard, O. S., Østergaard, L., ... & Agergaard, J. (2021, January). Persistent symptoms in hospitalized patients recovering from COVID-19 in Denmark. In Open Forum Infectious Diseases.
25. Bliddal, S., Banasik, K., Pedersen, O. B., Nissen, I., Cantwell, L., Schwinn, M., ... & Feldt-Rasmussen, U. (2021). Acute and persistent symptoms in non-hospitalized PCR-confirmed COVID-19 patients. medRxiv.
26. Petersen, M. S., Kristiansen, M. F., Hanusson, K. D., Danielsen, M. E., Gaini, S., Strøm, M., & Weihe, P. (2020). Long COVID in the Faroe Islands-a longitudinal study among non-hospitalized patients. Clinical Infectious Diseases.
27. Cirulli, E., Barrett, K. M. S., Riffle, S., Bolze, A., Neveux, I., Dabe, S., ... & Washington, N. L. (2020). Long-term COVID-19 symptoms in a large unselected population. medrxiv.
28. Jacobs, L. G., Gourna Paleoudis, E., Lesky-Di Bari, D., Nyirenda, T., Friedman, T., Gupta, A., ... & Aschner, J. L. (2020). Persistence of symptoms and quality of life at 35 days after hospitalization for COVID-19 infection. PloS one, 15(12), e0243882.
29. Clavario, P., De Marzo, V., Lotti, R., Barbara, C., Porcile, A., Russo, C., ... & Porto, I. (2020). Assessment of functional capacity with cardiopulmonary exercise testing in non-severe COVID-19 patients at three months follow-up. medRxiv.

30. Contreras-Andrade, R. I., Juárez-González, L. I., Arellano-Montellano, E. I., & Herrera-García, J. C. (2020). Persistencia de síntomas en pacientes después de la enfermedad por coronavirus (COVID-19) en un hospital de tercer nivel de Puebla, México. Medicina Interna de México, 36(6), 789-793.
31. Moradian, S. T., Parandeh, A., Khalili, R., & Karimi, L. (2020). Delayed Symptoms in Patients Recovered from COVID-19. Available at SSRN 3667840.
32. Savarraj, J. P., Burkett, A. B., Hinds, S. N., Paz, A. S., Assing, A. R., Juneja, S., ... & Choi, H. A. (2020). Three-month outcomes in hospitalized COVID-19 patients. medRxiv.
33. Sudre, C. H., Murray, B., Varsavsky, T., Graham, M. S., Penfold, R. S., Bowyer, R. C., ... & Steves, C. J. (2020). Attributes and predictors of Long-COVID: analysis of COVID cases and their symptoms collected by the Covid Symptoms Study App. medRxiv.
34. Townsend, L., Dowds, J., O'Brien, K., Sheill, G., Dyer, A. H., O'Kelly, B., ... & Bannan, C. Persistent Poor Health Post-COVID-19 Is Not Associated with Respiratory Complications or Initial Disease Severity. Annals of the American Thoracic Society, (ja).
35. Woo, M. S., Malsy, J., Pöttgen, J., Seddiq Zai, S., Ufer, F., Hadjilaou, A., ... & Friese, M. A. (2020). Frequent neurocognitive deficits after recovery from mild COVID-19. Brain communications, 2(2), fcaa205.
36. Chun, H. J., Coutavas, E., Pine, A., Lee, A. I., Yu, V., Shallow, M., ... & Kraft, B. D. (2021). Immuno-fibrotic drivers of impaired lung function in post-COVID-19 syndrome. medRxiv.
37. Horvath, L., Lim, J. W. J., Taylor, J. W., Saief, T., Stuart, R., Rimmer, J., & Michael, P. (2020). Smell and taste loss in COVID-19 patients: assessment outcomes in a Victorian population. Acta Oto-Laryngologica, 1-5.
38. Miyazato, Y., Morioka, S., Tsuzuki, S., Akashi, M., Osanai, Y., Tanaka, K., ... & Ohmagari, N. (2020, November). Prolonged and late-onset symptoms of coronavirus disease 2019. In Open forum infectious diseases (Vol. 7, No. 11, p. ofaa507). US: Oxford University Press.
39. Sykes, D. L., Holdsworth, L., Jawad, N., Gunasekera, P., Morice, A. H., & Crooks, M. G. (2021). Post-COVID-19 Symptom Burden: What is Long-COVID and How Should We Manage It?. Lung, 1-7.
40. Goërtz YMJ, Van Herck M, Delbressine JM, et al. Persistent symptoms 3 months after a SARS-CoV-2 infection: the post-COVID-19 syndrome? ERJ Open Research 2020;**6**(4):00542-2020 doi: 10.1183/23120541.00542-2020[published Online First: Epub Date]|
41. Garrigues, E., Janvier, P., Kherabi, Y., Le Bot, A., Hamon, A., Gouze, H., ... & Nguyen, Y. (2020). Post-discharge persistent symptoms and health-related quality of life after hospitalization for COVID-19. *Journal of Infection*, *81*(6), e4-e6.
