## Supplementary material for "Fatigue symptoms associated with COVID-19 in convalescent or recovered COVID-19 patients; a systematic review and meta-analysis": S-4

Supplementary material 4 (S-4). Additional analyses

| Study name | PCR | Fatigue Assessment | Time point (Days) | Statistics for each study: Event rate | Lower limit | Upper limit |
|---|---|---|---|---|---|---|
| Bliddal et al 2021 | + | Self-report | 14 | 0.162 | 0.117 | 0.220 |
| Cirulli et al 2020 | + | Self-report | 16 | 0.054 | 0.030 | 0.094 |
| | | | | 0.097 | 0.031 | 0.262 |

Event rate and 95% CI

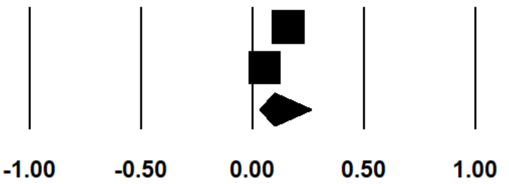


Figure S-4a. Prevalence of self-reported fatigue during the first month of COVID-19 recovery within samples recruited through patient registries or epidemiological datasets.

| Study name | PCR | Fatigue Assessment | Time point (Days) | Event rate | Lower limit | Upper limit |
|---|---|---|---|---|---|---|
| Bliddal et al 2021 | + | Self-report | 70 | 0.163 | 0.109 | 0.237 |
| Cirulli et al 2020 | + | Self-report | 76 | 0.080 | 0.042 | 0.146 |
| Horvath et al 2020 | + | Self-report | 69 | 0.029 | 0.010 | 0.087 |
| | | | | 0.082 | 0.034 | 0.188 |

Event rate and 95% CI

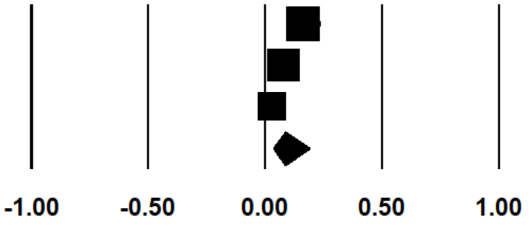


Figure S-4b. Prevalence of self-reported fatigue during the third month of COVID-19 recovery within samples recruited through patient registries or epidemiological datasets.

| Study name | Comparison | Time point (Days) | Statistics for each study: Risk ratio | Lower limit | Upper limit | Z-Value | p-Value |
|---|---|---|---|---|---|---|---|
| Woo et al. 2021 | Surivors vs. Controls | 85 | 4.053 | 0.230 | 71.379 | 0.956 | 0.339 |
| Cirulli et al 2020 | Surivors vs. Controls | 76 | 5.530 | 2.746 | 11.136 | 4.789 | 0.000 |
| Xiong et al 2020 | Surivors vs. Controls | 97 | 3.058 | 1.907 | 4.905 | 4.637 | 0.000 |
| | | | 3.688 | 2.502 | 5.436 | 6.592 | 0.000 |

Risk ratio and 95% CI

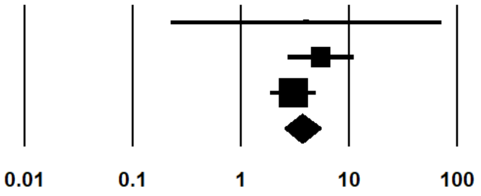


Figure S-4c. Fatigue incidence comparison between COVID-19 recovery and Healthy Controls during follow-up conducted 76 to 97 days post-recovery.

| Study name | Comparison | Time point (Days) | Statistics for each study | | | | |
|---|---|---|---|---|---|---|---|
| | | | Risk ratio | Lower limit | Upper limit | Z-Value | p-Value |
| Landi et al 2021 | PCR Neg vs. Pos | 47 | 1.028 | 0.651 | 1.621 | 0.117 | 0.907 |
| Su et al 2020 | PCR Neg vs. Pos | 30 | 0.599 | 0.341 | 1.052 | -1.783 | 0.075 |
| | | | 0.806 | 0.476 | 1.363 | -0.806 | 0.420 |

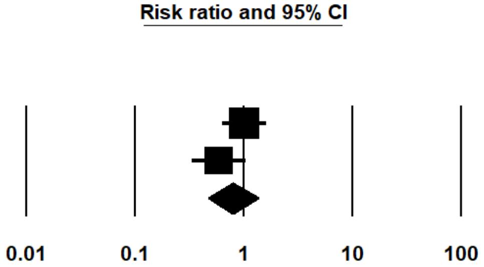


Figure S-4d. Fatigue incidence comparison between patients with negative PCR results and patients re-tested positive for COVID-19

| Study name | Comparison | Time point (Days) | Statistics for each study | | | | |
|---|---|---|---|---|---|---|---|
| | | | Odds ratio | Lower limit | Upper limit | Z-Value | p-Value |
| Bliddal et al. 2021 | Females vs. Males | 84 | 2.200 | 1.015 | 4.769 | 1.997 | 0.046 |
| Huang et al 2020 | Females vs. Males | 180 | 1.330 | 1.055 | 1.677 | 2.409 | 0.016 |
| Xiong et al 2020 | Females vs. Males | 97 | 1.887 | 1.280 | 2.782 | 3.205 | 0.001 |
| Sykes et al 2021 | Females vs. Males | 113 | 2.937 | 1.404 | 6.145 | 2.860 | 0.004 |
| | | | 1.782 | 1.273 | 2.495 | 3.366 | 0.001 |

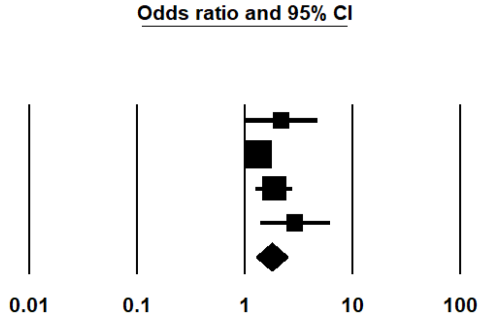


Figure S-4e. Self-reported Fatigue incidence comparison between Female and Male that recovered from COVID-19.

| Group by Report type | Study name | Comparison | Time point (Days) | Statistics for each study | | | | |
|---|---|---|---|---|---|---|---|---|
| | | | | Odds ratio | Lower limit | Upper limit | Z-Value | p-Value |
| Self-report | Bliddal et al. 2021 | Females vs. Males | 84 | 2.200 | 1.015 | 4.769 | 1.997 | 0.046 |
| Self-report | Huang et al 2020 | Females vs. Males | 180 | 1.330 | 1.055 | 1.677 | 2.409 | 0.016 |
| Self-report | Xiong et al 2020 | Females vs. Males | 97 | 1.887 | 1.280 | 2.782 | 3.205 | 0.001 |
| Self-report | Sykes et al 2021 | Females vs. Males | 113 | 2.937 | 1.404 | 6.145 | 2.860 | 0.004 |
| Self-report | | | | 1.904 | 1.124 | 3.226 | 2.394 | 0.017 |
| Validated | Venturelli et al. 2021 | Females vs. Males | 81 | 0.847 | 0.621 | 1.156 | -1.046 | 0.295 |
| Validated | Townsend et al 2020 | Females vs. Males | 72 | 2.867 | 1.478 | 5.559 | 3.117 | 0.002 |
| Validated | El Sayed et al 2020 | Females vs. Males | 12 | 0.596 | 0.358 | 0.993 | -1.988 | 0.047 |
| Validated | Halpin et al 2020 | Females vs. Males | 48 | 2.827 | 1.241 | 6.441 | 2.474 | 0.013 |
| Validated | | | | 1.285 | 0.751 | 2.199 | 0.915 | 0.360 |

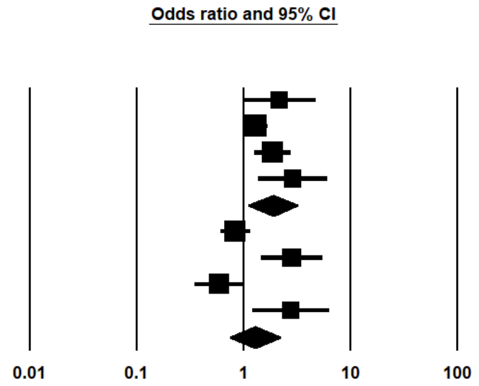


Figure S-4f. Gender differences in fatigue grouped by fatigue report type.

| Study name | Comparison | Time point (Days) | Odds ratio | Lower limit | Upper limit | Z-Value | p-Value |
|---|---|---|---|---|---|---|---|
| | | | Statistics for each study | | | | |
| Mandal et al 2020 | Severe vs. Non-severe | 54 | 1.592 | 0.765 | 3.310 | 1.244 | 0.214 |
| Xiong et al 2020 | Severe vs. Non-severe | 97 | 1.284 | 0.876 | 1.881 | 1.281 | 0.200 |
| | | | 1.344 | 0.958 | 1.886 | 1.711 | 0.087 |

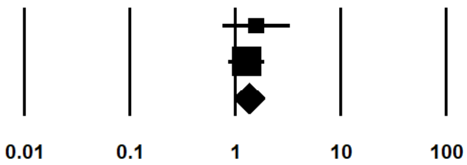


Figure S-4g. Self-reported Fatigue incidence comparison between recovery from severe and non-severe COVID-19.

| Study name | Comparison | Time point (Days) | Odds ratio | Lower limit | Upper limit | Z-Value | p-Value |
|---|---|---|---|---|---|---|---|
| | | | Statistics for each study | | | | |
| Halpin et al. 2020 | ICU vs. Ward | 48 | 1.683 | 0.677 | 4.185 | 1.120 | 0.263 |
| Townsend et al 2020 | ICU vs. Ward | 72 | 0.550 | 0.194 | 1.556 | -1.127 | 0.260 |
| | | | 0.991 | 0.332 | 2.960 | -0.017 | 0.987 |

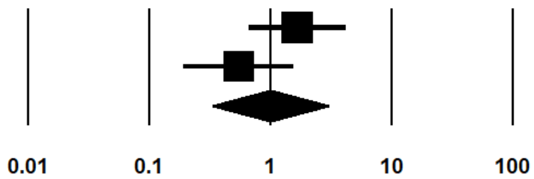


Figure S-4h. Fatigue incidence measured by validated scale between patients discharged from ICU and hospital Wards.

| Study name | Comparison | Time point (Days) | Odds ratio | Lower limit | Upper limit | Z-Value | p-Value |
|---|---|---|---|---|---|---|---|
| | | | Statistics for each study | | | | |
| Sykes et al 2020 | ICU vs. Ward | 113 | 0.716 | 0.295 | 1.740 | -0.738 | 0.461 |
| de Graaf et al. 2021 | ICU vs. Ward | 42 | 2.229 | 0.416 | 11.944 | 0.936 | 0.350 |
| Garrigues et al 2020 | ICU vs. Ward | 97 | 1.185 | 0.479 | 2.929 | 0.367 | 0.714 |
| | | | 1.024 | 0.566 | 1.853 | 0.078 | 0.938 |

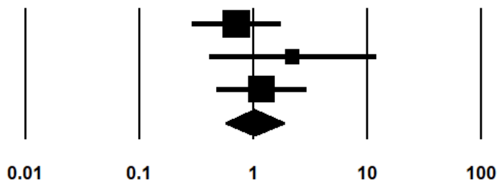


Figure S-4i. Self-reported Fatigue incidence comparison between patients discharged from ICU and hospital Wards.
