## Supplementary material for "Fatigue symptoms associated with COVID-19 in convalescent or recovered COVID-19 patients; a systematic review and meta-analysis": S-5

**Supplementary material 5 (S-5). Publication bias**

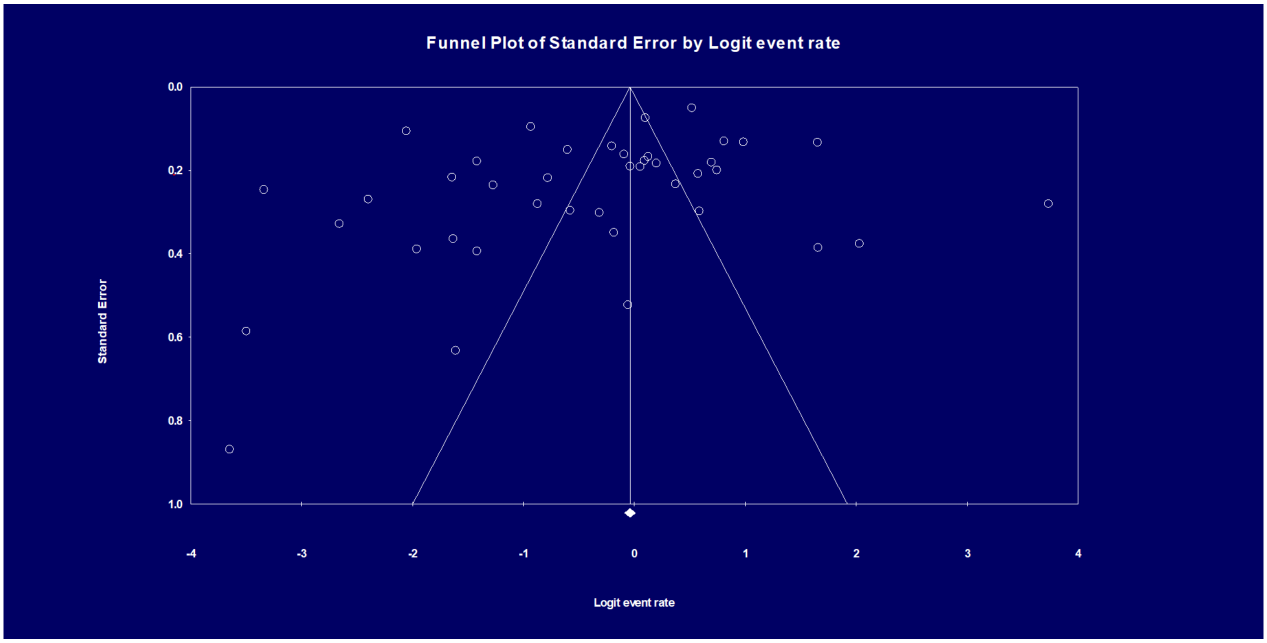


Figure S-2a. Funnel plot of studies associated with all included studies.

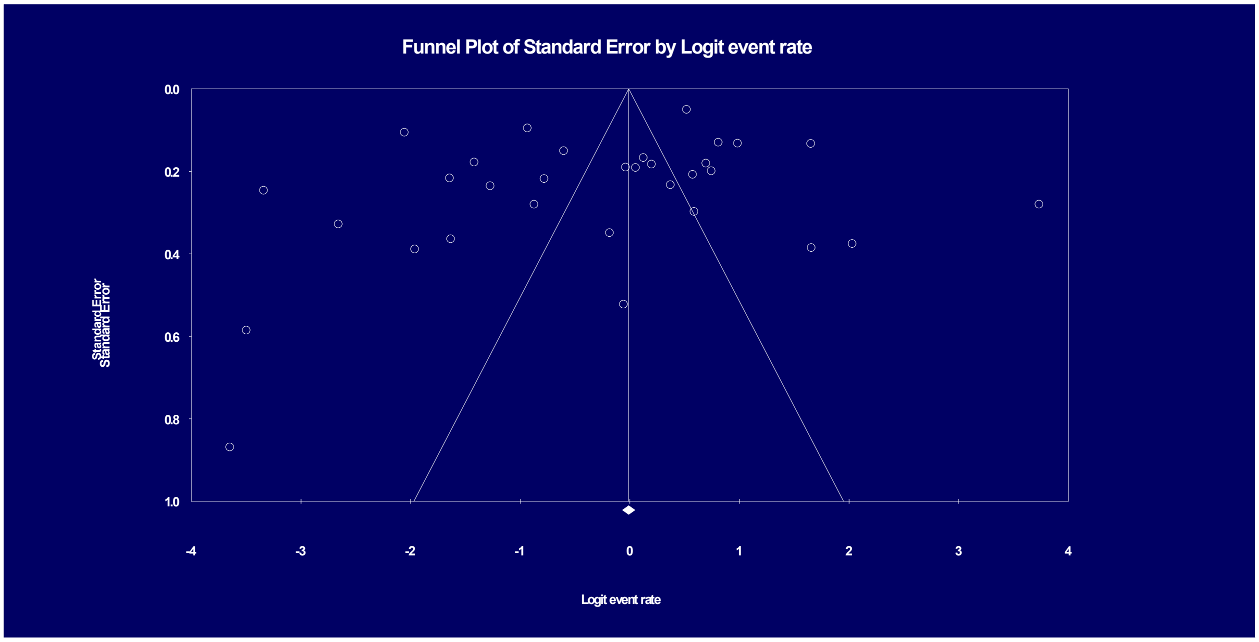


Figure S-2b. Funnel plot of studies in which fatigue was assessed through self-report.

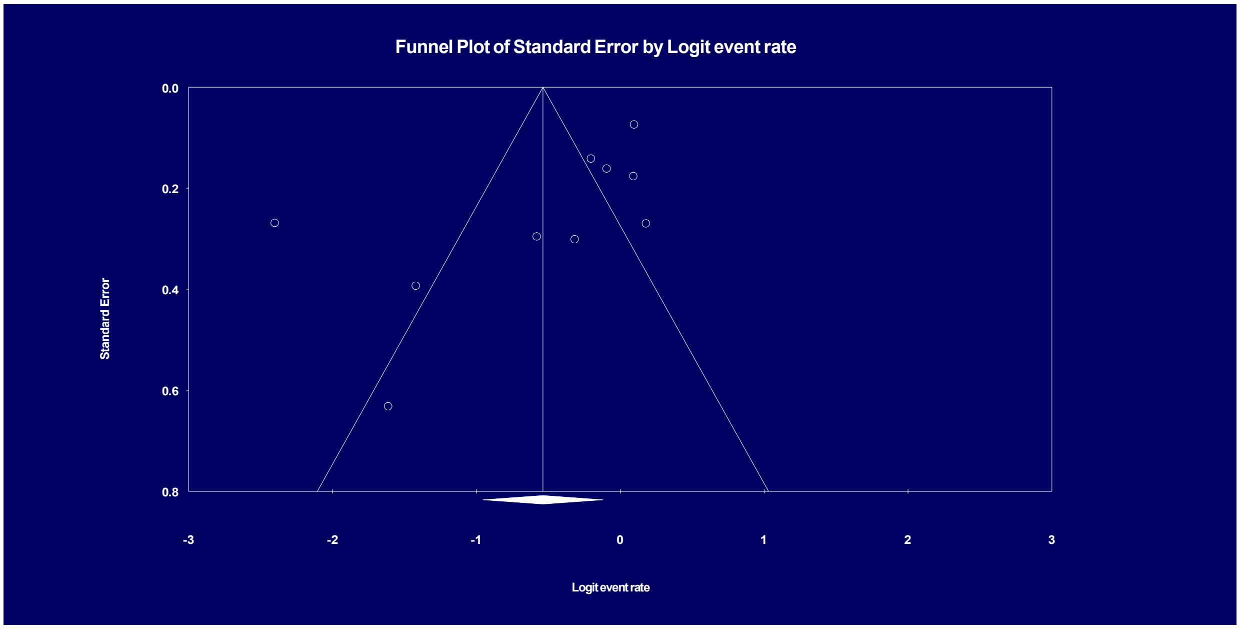


Figure S-2c. Funnel plot of studies in which fatigue was assessed through validated measures.
